# Single-cell profiling reveals venom-specific T-cells associated with tolerance during immunotherapy

**DOI:** 10.64898/2026.11.19.26368471

**Authors:** Florent Fauchère, Andreas Thiel, Julian Braun, Margitta Worm

## Abstract

**Background:** Venom immunotherapy (VIT) is well established, but the dynamics of the rare venom-specific T cells during treatment need to be investigated. Defining these responses may identify biomarkers and pathways associated with treatment-induced tolerance.

**Methods:** We sampled nine wasp-allergic individuals before and 7 days and 6 months after initiation of VIT, and 5 non-allergic donors at one time point. Single-cell RNA sequencing of isolated venom-specific T cells was used to characterise changes to immunophenotypes and immune receptor repertoires during VIT.

**Results:** We unveiled a broad phenotype diversity among venom-specific T cells, which were altered during VIT. Most notably, IL5⁺Th2 cells strongly correlated with Ves v 5-specific IgE levels at baseline, whereas their increase during VIT was associated with protective humoral immune responses, determined by the Ves v 5-specific IgG4/IgE ratio. Moreover, VIT induced exhaustion markers in Tconv, whereas Tregs acquired an activated phenotype, which was associated with increased specific IgG4 levels. T cell receptor (TCR) repertoire analysis revealed a highly polyclonal repertoire and substantial clonal expansion and plasticity in Tconv. By contrast, Treg clonotypes emerged during VIT, with no evidence of Tconv-to-Treg conversion.

**Conclusion:** VIT induces early phenotypic changes in venom-specific T cells, which are associated with a protective IgG4 response. In particular, IL5⁺Th2 and activated Tregs may represent candidate biomarkers of both antigen-specific sensitisation and VIT response and suggest a potential role of IL-5-related pathways in allergen-specific tolerance development. Furthermore, Tconv and Tregs TCR repertoires remained separated during VIT, opposing the Tconv-to-Treg transition hypothesis.

**Graphical Abstract,:** created in https://BioRender.com

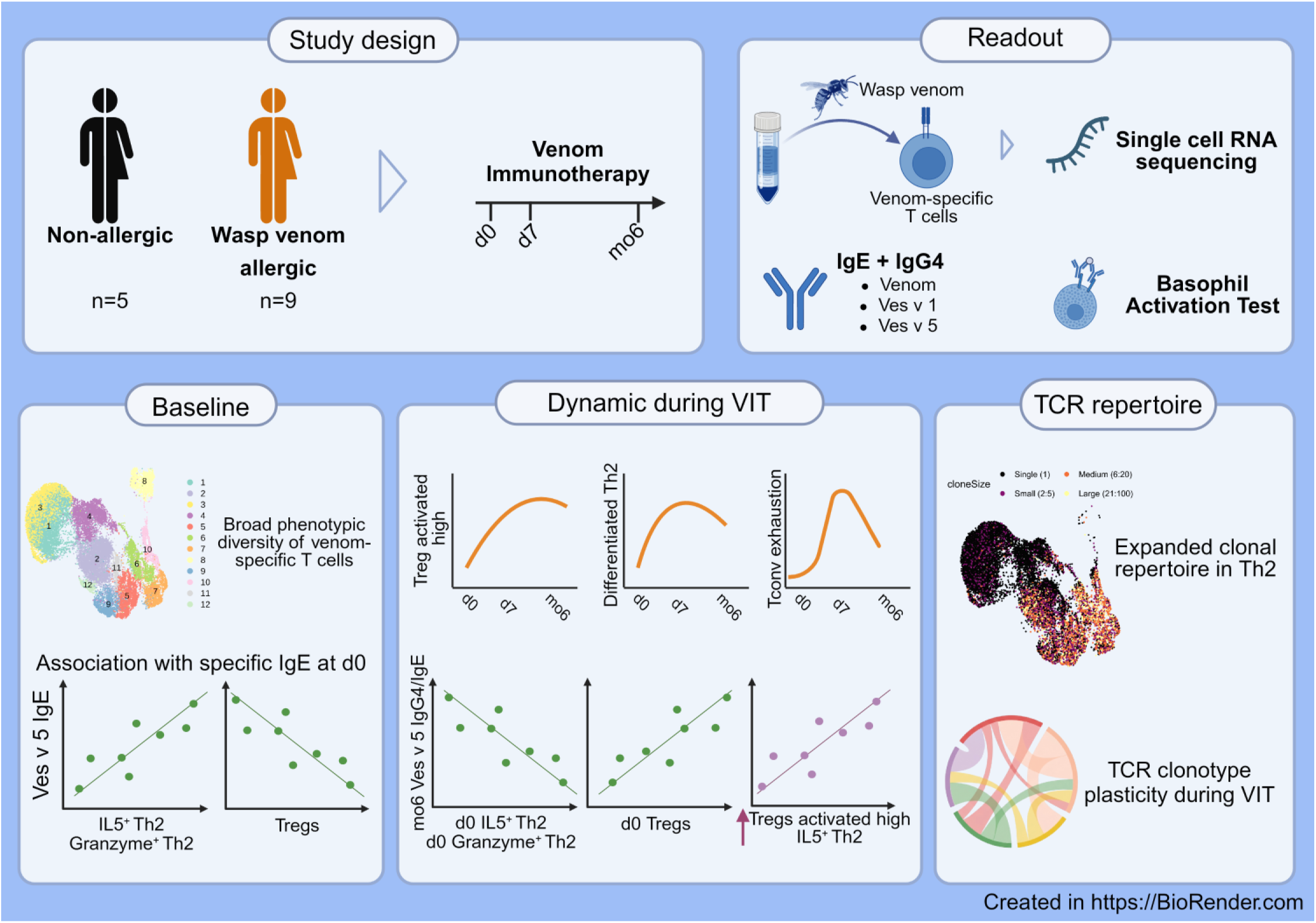

## 1. Introduction

Allergies are characterised by an exaggerated immune response to otherwise harmless antigens and can lead to life-threatening reactions. Until now, the only causal therapeutic approach is allergen-specific immunotherapy (AIT), aiming for long-term tolerance through repeated exposure to the allergen. Successful AIT is characterised by the transition of the type 2 immune response to a tolerant state with desensitisation of mast cells and basophils, an increase of allergen-specific IgG4 and Tregs and a decrease of allergen-specific Th2 cells [1], especially Th2A [2]. Nevertheless, the success of AIT differs to a major extent between allergens and individuals, and remission rates are allergen dependent. Therefore, an in-depth characterisation of the dynamics of allergen-specific lymphocytes, especially T and B cells, during AIT may unveil early biomarkers stratifying between responders and non-responders and identify immune pathways that could be targeted using immune modulatory treatments.

Insect venom allergy is one of the most frequent causes of anaphylaxis, particularly in adults [3,4]. Venom immunotherapy (VIT) is around 85% effective [5], rendering it a particularly useful setting to investigate mechanisms of tolerance induction in detail. Previous studies observed early dynamics of wasp venom-specific T cell responses after VIT initiation [6,7] but at lower phenotypic resolution and without investigating the late response and its association with the humoral immune response.

Therefore, we sampled 9 wasp-venom allergic individuals before, 7 days and 6 months after VIT initiation, along with 5 non-allergic controls, and characterised their venom-specific T cells using single-cell RNA sequencing (scRNAseq), as well as their humoral immune response. The data reveal 12 phenotypically distinct T cell populations among CD4 conventional T cells (Tconv) and regulatory T cells (Tregs) that were dynamically regulated during VIT. Specifically, the shift from one T cell population to another, as well as expression of genes related to exhaustion, were identified to correlate with the induction of specific IgG4. Furthermore, we used TCR sequencing to fate map T cell clones across multiple time points, revealing limited clonal expansion but high T cell plasticity, with new clonotypes arising in Tregs rather than contribution of already present Tconv.

## 2. Materials and Methods

### Participants and ethics

Eligible participants with wasp venom allergy and beginning VIT (n=10, one dropout) or no allergy (n=5) were recruited from the Division of Allergy and Immunology, Department of Dermatology, Venereology and Allergy, Charité, following the principles of the Declaration of Helsinki, and samples were collected before treatment (d0), 7 days (d7), and 6 months (mo6) after VIT initiation. Compared with the venom-allergic cohort, non-allergic individuals were younger and included a higher proportion of males (**Table S1**). VIT was performed using Venomil (Bencard Allergie GmbH, Munich, Germany) according to an ultra-rush protocol including dose escalation (day 1: 0.1, 1, 10µg; day 2: 10, 100µg, day 3: 100µg) followed by maintenance doses (100 µg weekly for 4 weeks and then every 4 weeks.). The study was approved by the local ethics committee (EA2/304/21).

### Sampling and cryo-banking of PBMCs

Blood was collected in Vacuette lithium heparin or in Vacuette serum tube (Greiner Bio-One, Monroe, U.S.). PBMC were isolated by gradient density centrifugation (Leucosep tubes, Greiner Bio-One, Monroe, NC; Biocoll, Bio&SELL, Feucht, Germany) and cryopreserved in liquid nitrogen at 2×10^7^ cells/ml in 45% 1640 medium (Gibco, Thermo Fisher Scientific, Waltham, U.S.), 45% FCS and 10% DMSO.

### Determination of serum antibody titers

Levels of total IgE and IgE specific for wasp venom extract and allergenic components Ves v 1 and Ves v 5 were determined in serum in collaboration with LaborBerlin. Specific and total IgG4 antibody levels in serum were analysed using the ImmunoCap Phadia 200 (Thermo Fisher Scientific, Waltham, U.S.). based on wasp venom extract and allergenic components Ves v 1 and Ves v 5. The detection of specific and total antibody levels included the additional application of several quality controls (low-, medium- and high-concentrated).

### Basophil Activation Test

80µl of whole blood was incubated at 37°C, 5% CO2 for 10 minutes before stimulation for 20 minutes with PBS, 10µg/ml anti-IgE antibodies (Goat polyclonal, Thermo Fisher Scientific, Waltham, U.S.), 10^-5M fMLP (Sigma-Aldrich, Burlington, U.S.), or 1ng/ml, 10ng/ml, 100ng/ml, 1µg/ml and 10µg/ml wasp venom extract i3 (DST-diagnostic, Schwerin, Germany). Staining antibodies are indicated in **Table S2**. Stimulations were stopped with 2nM EDTA, followed by fixation and permeabilisation using BD FACS lysing solution and BD Phosphoflow Perm Buffer II (BD Biosciences, Franklin Lakes, U.S.), and intracellular staining for 30 min. Samples were acquired using a MACSQuant^®^ Analyzer 16 (Miltenyi Biotech, Bergisch Gladbach, Germany) according to the gating strategy illustrated in **Figure S1**.

Two allergic participants were basophil non-responders at one or both time points, as defined by the absence of basophil activation following anti-IgE stimulation, and one sample could not be analysed because of technical issues. For each individual, CDMAX (highest percentage of CD63high basophils after stimulation with any concentration of allergen), EC50 (concentration of allergen in ng/ml required to reach 50% of the maximum percentage of CD63high basophils reached with allergen or anti-IgE stimulation), and the Area under the dose-response curve (obtained with allergen stimulation) were determined.

### Sorting of wasp venom-specific T cells

Cryopreserved PBMCs were rapidly thawed and washed twice with AIM-V medium (Thermo Fisher Scientific, Waltham, U.S.) containing 25U/ml Benzonase nuclease (Millipore, Sigma-Aldrich, Burlington, U.S.). B cells were enriched using CD19 microbeads and LS columns (Miltenyi Biotech, Bergisch Gladbach, Germany). The flow-through (CD19-) was used for T-cell stimulation and the retained cells (CD19+) for staining Ves v 5-specific B cells using dual-fluorochrome-labelled Ves v 5, although the results were omitted due to the absence of clear findings.

After resting for 2 hours at 37°C in AIM-V medium (Thermo Fisher Scientific, Waltham, U.S.), CD19-cells were washed with AIM-V supplemented with 25 U/ml benzonase nuclease (Sigma-Aldrich, Burlington, U.S.), and resuspended at 10^7^ cells/ml in AIM-V medium (Thermo Fisher Scientific, Waltham, U.S.) supplemented with 2.5% heat-inactivated AB serum (PAN-Biotech GmbH, Aidenbach, Germany), 1µl/ml CD28 antibodies (clone CD28.2, BD Biosciences, Franklin Lakes, U.S.) and CD40 antibodies (Miltenyi Biotech, Bergisch Gladbach, Germany). Samples were either stimulated with PBS (unstimulated control), 20µg/ml wasp venom extract (DST-diagnostic, Schwerin, Germany) or with 1.5 mg/ml SEB and 1 mg/ml TSST1 (Sigma-Aldrich, Burlington, U.S.). Incubation was performed at 37 °C, 5% CO2 for 16 hours.

Stimulation was stopped with 2 mM EDTA, staining was performed at room temperature for 30 minutes (**Table S3**), and the samples were fixed with DSP (Thermo Fisher Scientific, Waltham, U.S.), prepared as described by Attar et al. [8]. Fixation was stopped by addition of Tris HCL (final concentration 20mM). Samples were then enriched for CD137 and CD154 using anti-PE and anti-APC microbeads and MS columns (Miltenyi Biotech, Bergisch Gladbach, Germany). In addition, subsets of each sample were taken before enrichment to assess the percentage of activated T cells before enrichment. The enriched venom-specific T cells were sorted on a FACSDiscover S8 sorter (BD Biosciences, Franklin Lakes, U.S.) and identified as activated CD8 T cells (CD8+ CD137+), T conventional helper cells (CD4+ CD137+ CD154+) or as activated T reg (CD4+ CD137+ CD154-CD127-CD25+). The gating strategy is depicted in **Figure S2**.

### scRNAseq library preparation and sequencing

Sorted cells were merged, centrifuged and resuspended in PBS + 0.05%BSA at 500 cells/µl. For each of the 5 batches, libraries for gene expression, TCR, and cell surface proteins were prepared using the v3 GEM-X 5’ (10X Genomics, Pleasanton, U.S.) reagents and workflow. Libraries were sequenced on a NovaSeq X Plus 10B-100 cycles (Illumina Inc., San Diego, U.S.) and demultiplexed using the Cell Ranger Multi workflow (10X Cloud).

### scRNAseq analysis

scRNAseq data were analysed with Seurat v5 [9]. After quality control, 38,606 cells from 37 samples were retained for analysis (**Figures S3a,b**). Datasets were integrated using SCTransform [10] and Harmony [11], resulting in successful batch correction (**Figure S3c**). Unsupervised clustering using Leiden algorithm [12] identified 12 cell populations labelled using lineage marker expression (**Figures 3e,f**). Two low-quality T-cell clusters, characterised by low RNA content, reduced CD3E expression, increased mitochondrial transcripts, and absence of TCR sequences, were excluded. The remaining cells were separated into B-cell and T-cell datasets for subsequent analyses, including re-integration and clustering. Gene set enrichment analysis (GSEA) were performed using the fgsea package [13] and the Reactome gene set [14] provided by the msigdbr package [15].

### scAIRR-seq analysis

Single-cell adaptive immune receptor repertoire sequencing (scAIRR-seq) from T cells was integrated using scRepertoire in R [16]. The expansion and transition of TCR clones across time points and in T cell clusters were assessed using STARTRAC [17]

### Data processing and statistical analysis

Clinical data were collected and managed using REDCap electronic data capture tools hosted at Charité. All statistical analyses were calculated using R (version 4.5.2, R Foundation, Vienna, Austria) and RStudio (version 2024.04.1+748, Posit PBC, Boston, U.S.). Since the data were non-normally distributed, non-parametric comparisons were performed (paired or unpaired Wilcoxon tests and Spearman correlation test) and were adjusted for multiple comparisons between groups or time points using Benjamini-Hochberg (FDR implementation). Results of statistical comparisons are displayed as p-values.

## Data Availability

All data produced in the present study are available upon reasonable request to the authors.

## Abbreviations

(AIT): Allergen immunotherapy
(VIT): Venom immunotherapy
(scRNAseq): Single-cell RNA sequencing
(Treg): Regulatory T cell
(Tconv): CD4 conventional T cell
(BAT): Basophil activation test
(TCR): T cell receptor
(BCR): B cell receptor
(sIgE): specific IgE
(sIgG4): specific IgG4

## Data availability

All R scripts used for data analysis are publicly available on GitHub: https://github.com/faucherf/AGThiel_AGWorm_VIT-WV-Tcell-scRNAseq

## Acknowledgements and Funding

This work was funded by the Deutsche Forschungsgemeinschaft (DFG, German Research Foundation) as part of the clinical research unit (CRU339): Food allergy and tolerance (FOOD@) – 409525714.

We thank Dr. Sabine Dölle-Bierke, Lea Katharina Faust, Elsbeth Oestmann and Josefine Grünhagen from the Division of Allergy and Immunology, Department of Dermatology, Venereology and Allergology, Charité – Universitätmedizin, Berlin for supervising the clinical management, clinical data and sample collection.

We thank Anja Alfandega from the Klinik für Pädiatrie m.S. Pneumologie, Immunologie und Intensivmedizin for the serological measurements, and the Genomics Technology Platform from the Berlin Institute of Health for the sequencing.

We thank Louis Helmut Löser for the help with the processing of the samples.

## Author contributions

F.F. planned and performed experiments, analysed the data and wrote the manuscript. J.B.,

M.W. and A.T. designed and supervised the study.

## Competing interests

All authors declare no competing interests.

## Results

### Comprehensive profiling reveals a broad phenotypic diversity of venom-specific T cells

Venom-specific T cells were identified after *in vitro* stimulation with venom extract and magnetic enrichment, and sorted as venom-specific CD4 Tconv, Treg or CD8 T cells (**Figure 1a, S2**). We detected very few venom-specific CD8 T cells, hindering a robust downstream analysis (**Figure S6c**). In contrast, sufficient venom-specific Tconv and Treg were detected in allergic individuals, with substantial inter-individual variability, and remained stable during VIT (**Figures S6d,e**).

**Figure 1:**
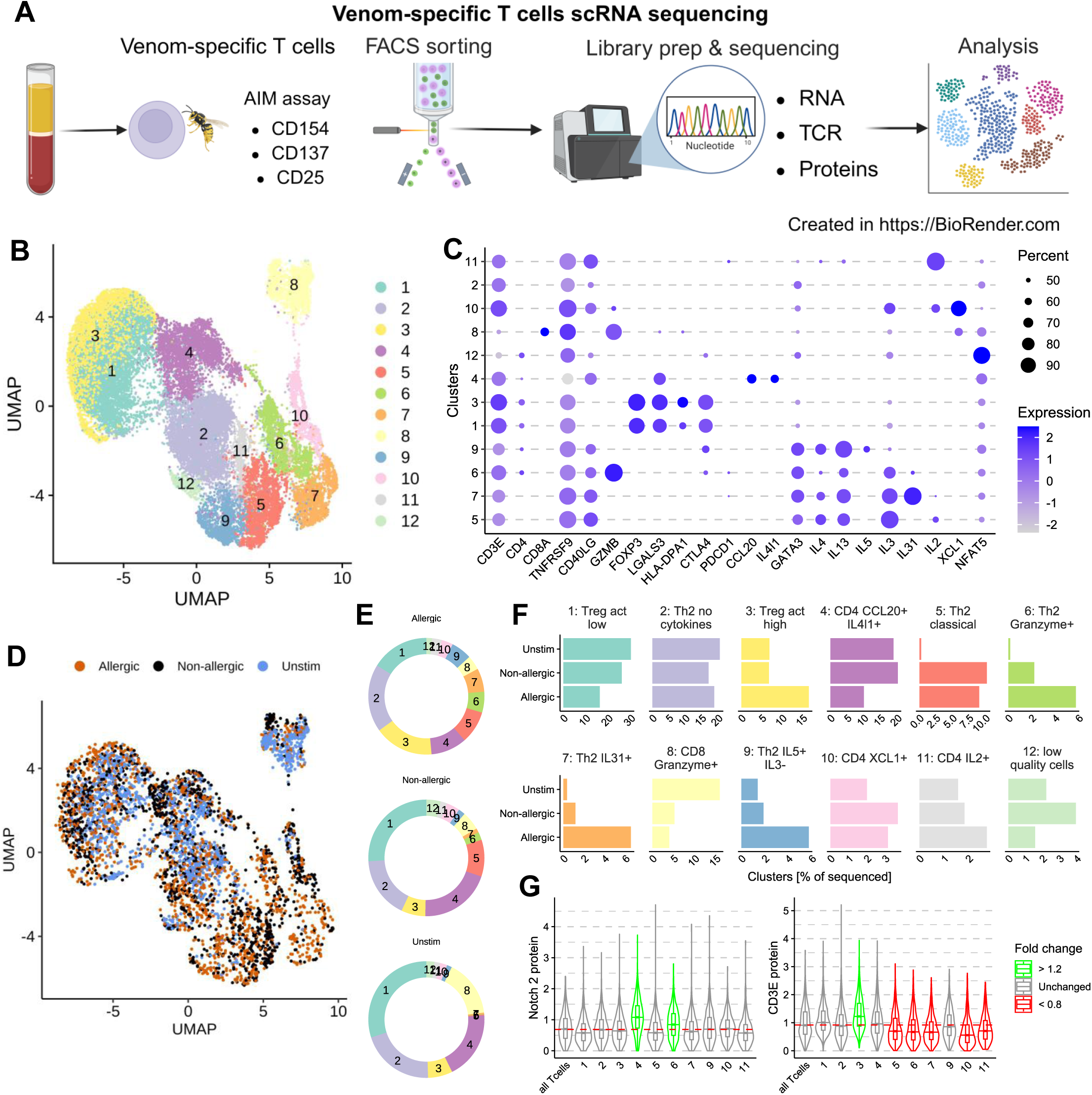
Venom-specific T cell profiling reveals a broad range of T cell responses. **A:** Scheme depicting the experimental setup, created in https://BioRender.com. **B:** UMAP representing venom-specific T cells with Leiden clusters as colours. **C:** Heatmap depicting selected gene expression in T cell clusters, with gene expression level as colours and percentage of cells expressing the gene as size. **D:** UMAP with venom-allergic, non-allergic or unstimulated samples as colours. **E:** Doughnut plots and **F:** Bar plots depicting percentages of T cell clusters in allergic, non-allergic or unstimulated samples. **G:** Violin plot depicting Notch 2 or CD3E protein expression in venom-specific T cells.

The sorted T cells were measured by scRNAseq and the filtered dataset was integrated and corrected for batch effects (**Figure S4a**), and 12 clusters were identified (**Figure 1b, Table S4**). Clusters were manually annotated based on cluster-specific differentially expressed genes (**Figure S4f**), selected marker genes (**Figure 1c; S4g**), and reactome gene set enrichment pathway (**Figure S5**). To distinguish antigen-specific from background populations, we also sorted activated cells from unstimulated samples and compared their cluster frequencies with those of venom-stimulated in venom-allergic and non-allergic (**Figure 1d–f**). In addition, surface expression of CD3 and Notch 2 was quantified using oligonucleotide-conjugated antibodies, as transcript abundance did not accurately reflect protein expression (**Figure 1g**, **Figure S4b-e)**. This was particularly relevant for CD3, which we previously identified as a surrogate marker of high functional avidity in antigen-specific conventional T cells [18]. Notch 2 was included because of its reported but controversial roles in immune tolerance and allergic inflammation in murine models [19–21].

Cluster 8 presented a cytotoxic CD8 phenotype, expressing CD8 and GZMB. Cluster 12 lacked a distinct transcriptional signature but expressed NFAT5, indicative of cellular stress. Both clusters were predominantly detected in unstimulated or non-allergic samples, thus unlikely to be venom-specific and were excluded from downstream analyses.

Clusters 1 and 3 expressed canonical Treg markers (FOXP3, TNFRSF9, IKZF2, LTB). Cluster 3 was most abundant in allergic samples, expressed higher CD3 protein levels, and upregulated activation-associated genes, including LGALS3, CD70, HLA-DPA1/DPB1, and HLA-DRA (**Figure S4h**), indicating stronger activation and potentially a more suppressive phenotype. These clusters were therefore annotated as activated-low and activated-high Tregs, respectively.

Clusters 5, 6, 7 and 9 strongly expressed Th2-associated genes GATA3, IL4 and IL13, and were absent from unstimulated samples. Clusters 5–7 downregulated CD3 protein, consistent with high functional avidity, whereas IL5⁺IL3⁻ cluster 9 retained CD3 expression despite a terminally differentiated phenotype [22]. Th2 cluster 6 and cluster 7 were further distinguished by expression of GZMB and Notch 2 or IL31.

Cluster 2 expressed GATA3 and CD40LG but lacked Th2 cytokines, suggesting an anergic or tolerogenic phenotype. This was supported by GSEA, revealing expression of genes involved in the GPCR pathways and genes involved in anti-inflammatory response favouring parasite infection (**Figure S5**). However, its comparable abundance across stimulations suggests background activation.

We could not assign a definitive identity to clusters 4, 10, and 11. Cluster 4 expressed CCL20, IL4I1, and LGALS3, displayed the highest Notch 2 protein expression, and was enriched in unstimulated and non-allergic, suggesting an activated population with migratory or immunoregulatory properties. Cluster 10 was characterised by high XCL1 expression and low CD3 protein, suggesting a role in supporting cross-presentation by CD8 T cells or NK cells. Cluster 11 was more enriched in venom-allergic, expressed IL2 and downregulated CD3 protein, and could support the proliferation of activated T cells.

### Venom-specific T-cell signatures are associated with allergic status and specific IgE

We next determined the composition of venom-specific CD4 T-cell populations in non-allergic and venom-allergic before VIT. Cluster frequencies were calculated relative to their parent populations, with Treg clusters (1 and 3) expressed as percentage of activated Tregs and conventional CD4 T-cell clusters (2, 4, 5, 6, 7, 9, 10, and 11) as percentage of activated CD4 Tconv cells.

As expected, venom-allergic presented higher levels of venom- and Ves V 5-specific IgE than non-allergic individuals (**Figure S6a**). The frequencies of venom-specific Tregs, cytokine-negative Th2 cells, classical Th2 cells (IL4⁺IL13⁺), IL2⁺ T cells, and CCL20⁺IL4I1⁺ T cells were comparable between venom-allergic and non-allergic. In contrast, terminally differentiated IL5⁺IL3⁻ Th2 cells, IL31⁺ Th2 cells, granzyme-expressing Th2 cells, and XCL1⁺ T cells were enriched in venom-allergic (**Figure 2a**).

**Figure 2:**
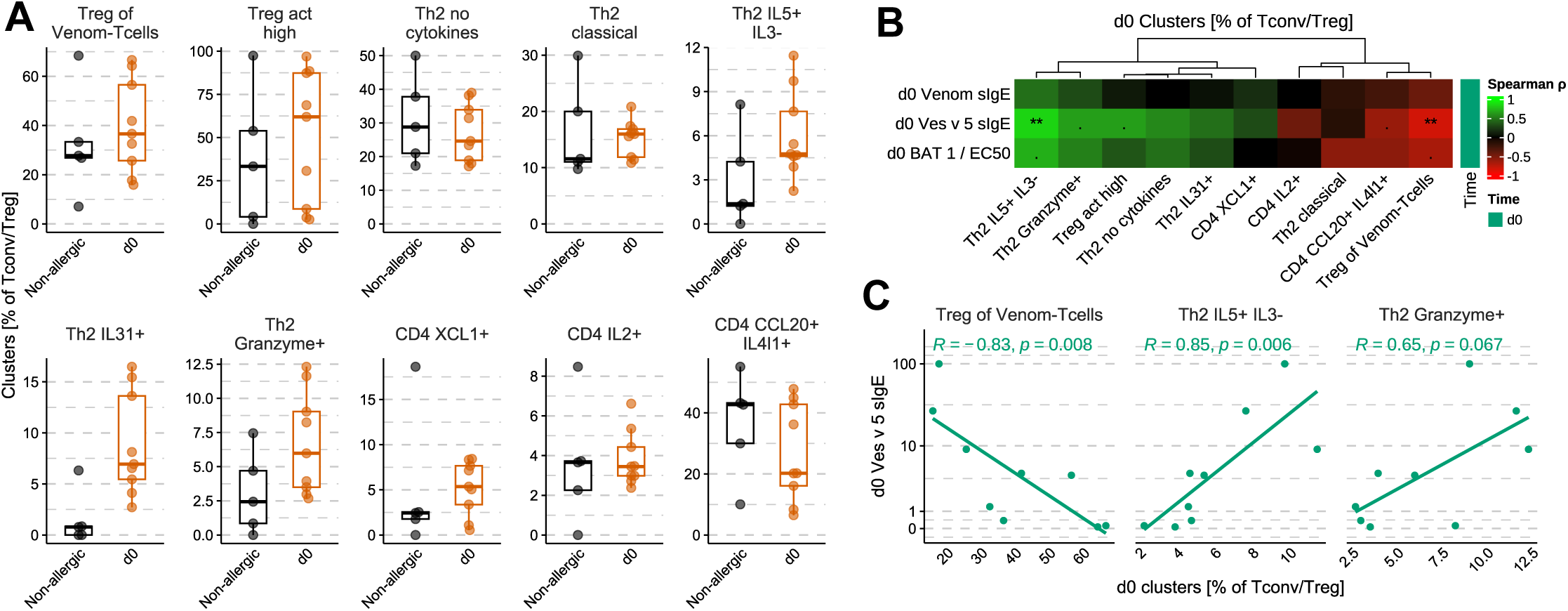
Venom-specific T cell signatures are associated with allergic status and specific IgE. **A:** Boxplots depicting percentages of T cell populations in non-allergic (black) or venom-allergic (orange) at d0. **B:** Heatmap presenting all, and **C:** plots presenting selected Spearman correlations between T cell population percentages and Venom sIgE, Ves v 5 sIgE or 1/EC50 (BAT) in venom-allergic individuals at d0.

Next, we examined the association of venom-specific T-cell subsets with the allergen-specific humoral immune response in venom-allergic individuals at baseline. Ves v 5-specific IgE correlated positively and strongly with IL5⁺IL3⁻ Th2 cells, but weakly with granzyme-expressing Th2 cells and activated-high Tregs (**Figure 2b,c; S7a**). Conversely, the frequency of total venom-specific Tregs correlated negatively with Ves v 5-specific IgE. Similar trends were observed for wasp venom-specific IgE and basophil sensitivity (1/EC50), although these correlations did not reach statistical significance because of greater inter-individual variability. Finally, the frequency of venom-specific CD4 Tconv measured by flow cytometry strongly correlated with Ves v 5-specific IgE (**Figure S7b**).

### Early serological VIT responses are detectable one week after initiation

To assess the humoral immune response during VIT, we measured serum-specific IgE (sIgE) and IgG4 (sIgG4) against venom, Ves v 1, and Ves v 5, but also performed basophil activation tests (BAT) to assess their functional impact. In most venom-allergic, sIgE levels transiently increased at d7 before returning to baseline by mo6 (**Figure 3a**). In contrast, sIgG4 levels against wasp venom and Ves v 5 increased by d7 and remained elevated until mo6 in most participants (**Figure 3b**). Consequently, the sIgG4/sIgE ratio increased at mo6 for both venom and, more prominently, Ves v 5 (**Figure 3c**). Total IgE concentrations remained stable, indicating that the observed changes were antigen-specific (**Figure S6b**).

**Figure 3:**
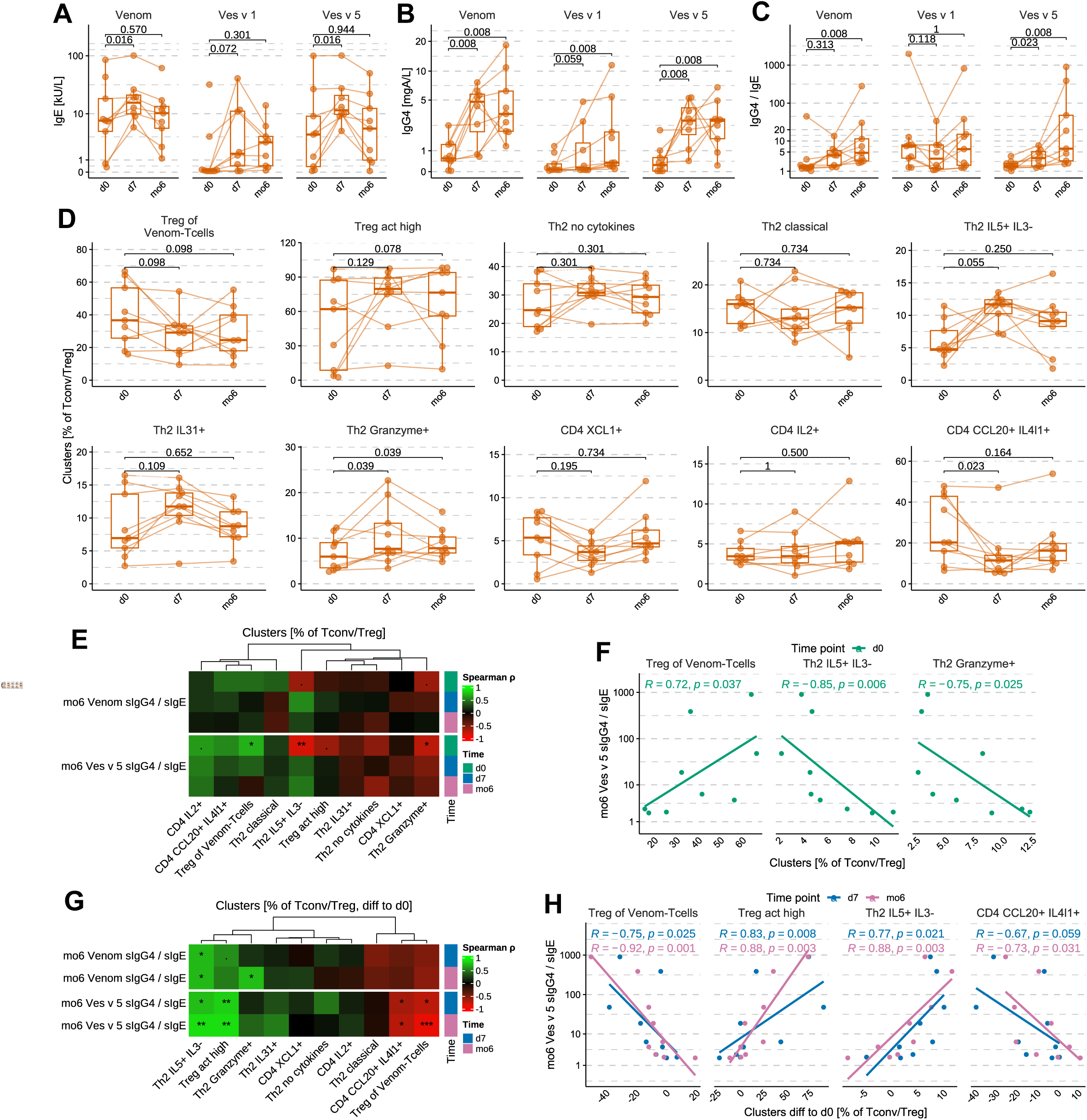
Venom-specific T cell dynamics are associated with the humoral immune response. Boxplots depicting biomarkers in venom-allergic (orange, n=9) at d0, d7 and mo6 for**: A:** Venom, Ves v 1 and Ves v 5-specific IgE, **B:** Venom, Ves v 1 and Ves v 5-specific IgG4, **C:** Venom, Ves v 1 and Ves v 5-specific IgG4/IgE ratio. **D:** Boxplots depicting percentages of T cell populations in allergic individuals at d0, d7 and mo6. Statistical testing was conducted using the Wilcoxon signed-rank paired test corrected for multiple comparisons between time points with Benjamini-Hochberg (FDR). **E:** Heatmap presenting all, and **F:** plots presenting selected Spearman correlations between T cell population percentages at d0, d7 or mo6 and Venom or Ves v 5 sIgG4/sIgE at mo6. **G:** Heatmap presenting all, and **H:** plots presenting selected Spearman correlations between T cell population percentages normalised to d0 at d7 or mo6 and Venom or Ves v 5 sIgG4/sIgE at mo6.

BAT was performed at d0 and mo6 and activated basophils were defined as CD63high (**Figure S1**). All non-allergic controls had negative BAT results. Four of the six venom-allergic showed an increase in EC50 after 6 months of VIT, indicating the functional relevance of their humoral immune modulation (**Figure S6f**). Nevertheless, we were unable to identify a clear trend in alternative BAT readouts, i.e. area under the curve or CDMAX (**Figure S6g,h**).

Together, our findings reproduce the humoral effectiveness of VIT. The sIgG4/sIgE ratio, particularly for Ves v 5, was therefore used as a marker of therapy success [5,23,24] in the following analysis.

### Venom-specific T cell dynamics are associated with the humoral immune response

To delineate the T cell subtype repertoire upon VIT, we next investigated the dynamics of venom-specific CD4 T-cell subsets. The data revealed heterogeneous changes among the analysed individuals and T cell populations (**Figure 3d**). The overall proportion of venom-specific Tregs showed a slight decrease at d7 and mo6, whereas the activated-high Treg subset expanded markedly at d7 and remained stable or declined only modestly by mo6. Cytokine-negative Th2 cells transiently increased at d7, while classical Th2 and XCL1⁺ T cells showed the opposite trend. In contrast, IL5⁺IL3⁻ Th2 cells increased in all but one patient at d7 and remained elevated at mo6 in most individuals. IL31⁺ and granzyme⁺ Th2 subsets also expanded early during VIT but returned towards baseline by mo6. Finally, the CCL20⁺IL4I1⁺ T-cell population decreased at d7 and remained low throughout follow-up in all but one participant.

To determine whether the T cell composition may support predicting treatment outcome, we correlated the frequencies of each T cell subset with the venom- and Ves v 5-specific IgG4/IgE ratios at mo6 (**Figure 3e, f, S7c**). Higher baseline frequencies of IL5⁺IL3⁻ and granzyme⁺ Th2 cells were associated with lower Ves v 5-specific IgG4/IgE ratios, as the subsets represent highly differentiated allergen-specific Th2 cells. Conversely, higher baseline frequencies of venom-specific Tregs correlated with higher Ves v 5-specific IgG4/IgE ratios, suggesting that a larger pre-existing pool of allergen-specific Tregs favours successful desensitisation.

T-cell subset frequencies at d7 or mo6 were not significantly associated with the IgG4/IgE ratio, and thus we assessed whether changes in T cell subsets during VIT, normalised to baseline, were associated with treatment response (**Figure 3g,h S7d**). Relative expansion of IL5⁺IL3⁻ Th2 cells positively correlated with the venom- and Ves v 5-specific IgG4/IgE ratios, likely reflecting the greater fold change in individuals with lower baseline frequencies. In contrast, expansion of the total venom-specific Treg compartment negatively correlated with the IgG4/IgE ratio, whereas increasing activated-high Treg subset among venom-specific Tregs was strongly positively correlated. These findings suggest that the differentiation towards an activated regulatory phenotype, rather than expansion of the overall Treg pool, is associated with successful VIT. Changes in the CCL20⁺IL-4I1⁺ population inversely correlated with the IgG4/IgE ratio, although this association may reflect the relative expansion of other T cell subsets.

### Transient T cell exhaustion is observed during VIT

We next assessed *in vivo* T cell activation by measuring CD38, HLA-DR, and MKI67, which are expected to be upregulated with delayed kinetics following stimulation and therefore should not be substantially induced during our 16-h *in vitro* stimulation. None of these markers showed strong expression across time points or T cell clusters (**Figure S8a**), suggesting limited *in vivo* T cell activation or proliferation.

We then evaluated T-cell exhaustion using a selected gene set (CTLA4, LAG3, PDCD1 and TIGIT). Treg clusters (1 and 3) were excluded due to constitutive expression of these genes.

As the expression dynamics of individual exhaustion markers differed across T cell subsets (**Figure S8b**), we calculated an exhaustion score by scaling and aggregating the expression of the four genes.

Except for the CCL20⁺IL4I1⁺ subset, all T cell populations showed an increased exhaustion score at d7 following VIT (**Figure 4a**). This increase was most pronounced in IL5⁺IL3⁻ Th2 cells and was largely driven by CTLA4 expression (**Figure S8b**). In most subsets, exhaustion scores declined by mo6 but remained above baseline, particularly in granzyme⁺ Th2 and XCL1⁺ T cells. However, we cannot exclude that prolonged exhaustion prevents reactivation of a fraction of venom-specific T cells upon *in vitro* stimulation, limiting reactivation and subsequent detection of venom-specific T cells by our activation-based enrichment strategy.

**Figure 4:**
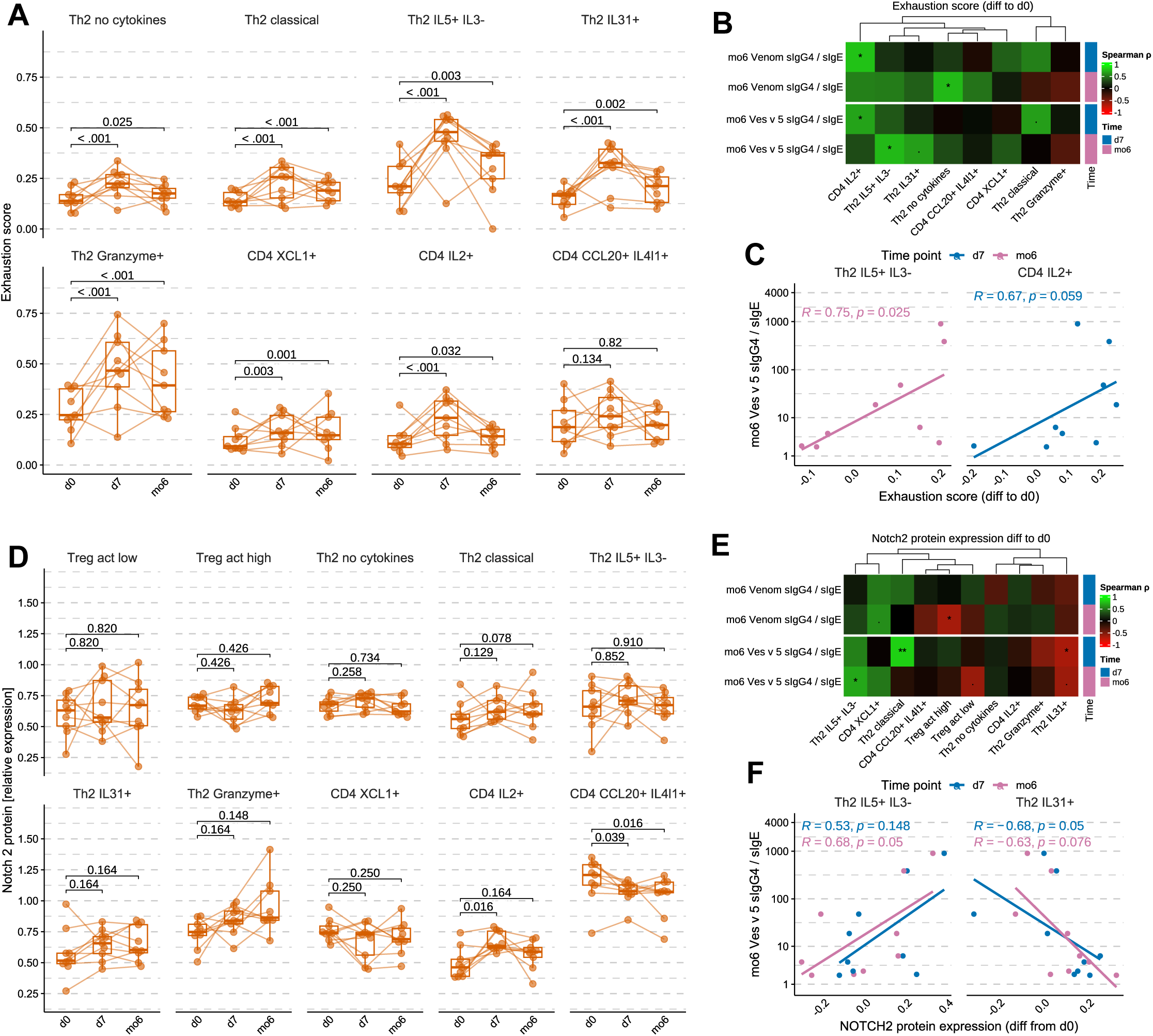
VIT induces transient expression of exhaustion markers in venom-specific T cells. **A:** Boxplots depicting exhaustion composite score in T cell populations in allergic individuals at d0, d7 and mo6. Statistical testing was conducted using the Wilcoxon signed-rank paired test corrected for multiple comparisons between time points with Benjamini-Hochberg (FDR). **B:** Heatmap presenting all, and **C:** plots presenting selected Spearman correlations between T cell population exhaustion score normalised to d0 at d7 or mo6 and Venom or Ves v 5 sIgG4/sIgE at mo6. **D:** Boxplots depicting Notch 2 protein expression in T cell populations in allergic individuals at d0, d7 and mo6. Statistical testing was conducted using the Wilcoxon signed-rank paired test corrected for multiple comparisons between time points with Benjamini-Hochberg (FDR). **E:** Heatmap presenting all, and **F:** plots presenting selected Spearman correlations between T cell population Notch 2 protein expression normalised to d0 at d7 or mo6 and Venom or Ves v 5 sIgG4/sIgE at mo6.

Correlation analyses revealed weak to moderate positive associations between exhaustion scores at d7 or mo6 and the venom- and Ves v 5-specific IgG4/IgE ratios at mo6 in most subsets, particularly in IL5⁺IL3⁻ Th2 cells at mo6 (**Figure 4b,c, S8c**).

### Notch 2 protein expression is associated with IgG4 induction

Given the debated role of Notch 2 in allergy, we assessed its protein expression during VIT using oligonucleotide-tagged antibodies. Notch 2 expression showed heterogeneous dynamics across allergic individuals, especially within IL2^+^ Tconv cells (**Figure 4d**). Its expression correlated positively with the specific IgG4/IgE ratio in classical Th2 cells at d7 and in IL5⁺IL3⁻ Th2 cells at mo6, while an inverse correlation was observed in IL31⁺ Th2 cells (**Figure 4e,f; S8d**). These associations support a potential role for Notch 2 in VIT-induced tolerance.

### TCR sequencing reveals a diverse repertoire with high phenotypic plasticity in Tconv and the emergence of new clonotypes in Treg during VIT

We next characterised the TCR repertoire of venom-specific T cells. Samples from the first batch were excluded because of low TCR recovery (**Figure S9a**). Clonal size and frequency analysis revealed greater expansion in Th2 clusters and the XCL1⁺ population, whereas Tregs and CCL20⁺IL4I1⁺ and IL2⁺ T cells showed limited clonal expansion (**Figure 5a; S9b**). Overall, the normalised Shannon entropy indicated a diverse TCR repertoire, although Tconv populations showed reduced diversity in individual donors (**Figure 5b**). Clonotype analysis further showed that Treg clones were largely restricted to Treg clusters, whereas Tconv clonotypes were frequently shared across multiple Tconv clusters (**Figure 5c, S9c** separated by individuals). Furthermore, TCR repertoires showed overlap across time points, but not between individuals (**Figure S10**).

**Figure 5:**
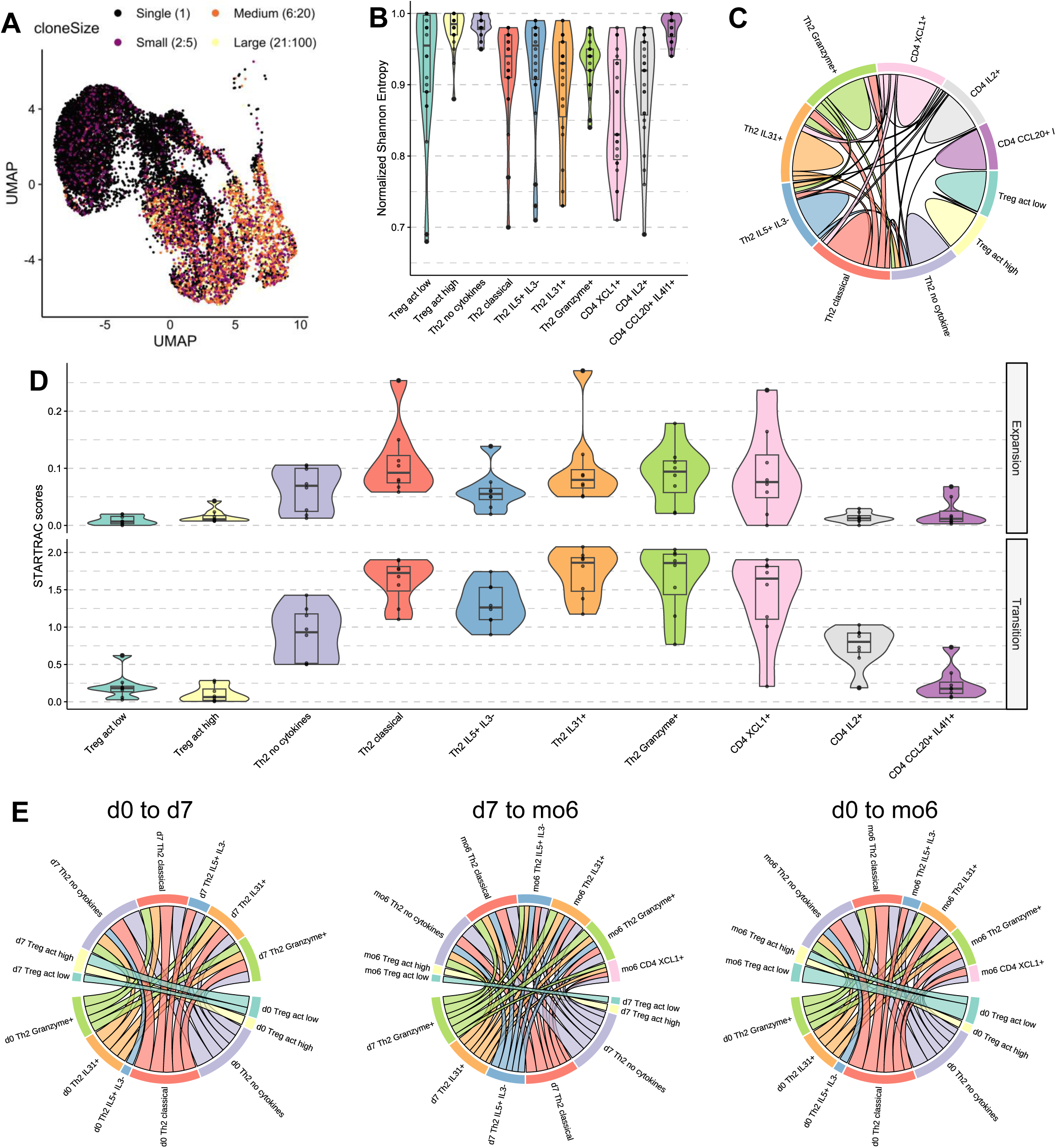
TCR sequencing reveals a diverse repertoire with high phenotypic plasticity in Tconv and the emergence of new clonotypes in Treg during VIT. **A:** UMAP illustrating clone size for each venom-specific T cell. **B:** Normalised Shannon entropy in each T cell population in venom-allergic (n=6 at each 3 time points). **C:** Circos plot representing clonotypes shared by multiple T cell populations at d0. **D:** Expansion and transition score of clonotype in T cell population during VIT. **E:** Circos plots depicting clonotype transition across T cell populations from d0 to d7, d7 to mo6 and d0 to mo6.

We next assessed clonotype expansion and transitions between time points using STARTRAC [17]. This analysis revealed limited clonal expansion but high transition scores in Tconv, suggesting that clonotypes shifted between phenotypic populations during VIT (**Figure 5d**). To characterise these transitions in greater detail, we tracked clonotypes across time points and T-cell populations (**Figures 5e, S11a-c** separated by individuals). Treg clonotypes assigned to the low-activation cluster at d0 predominantly transitioned to the high-activation cluster at d7 and remained in this state at mo6, consistent with progressive Treg activation during VIT. IL5⁺IL3^-^Th2 clonotypes at d0 were associated with cytokine-negative Th2 cells at d7, consistent with an anergic state, whereas classical and cytokine-negative Th2 cells at d0 were associated with IL5⁺IL3^-^ Th2 cells at d7, suggesting further differentiation. Greater plasticity was observed between d7 and mo6, potentially reflecting the longer time available for phenotypic transition. Other Th2 populations showed transitions across all time points, suggesting more dynamic phenotypes that may be shaped by the context and frequency of antigen encounters rather than stable cellular imprinting.

Finally, we assessed the dynamics of the top clonotypes within the Tconv and Treg compartments for each individual (**Figure S12a,b**). In Tconv cells, most dominant clonotypes detected at d0 persisted at d7 and mo6, with only a few individuals showing clonotype loss or generation between d0 and d7. In contrast, only a subset of Treg clonotypes persisted across time points, suggesting that VIT promotes the emergence of new venom-specific Treg clonotypes.

Overall, the TCR repertoire was diverse and not restricted to individual T-cell populations and during VIT, Tconv clonotypes persisted across time points with marked phenotypic plasticity, whereas Treg clonotypes showed limited plasticity with the emergence of new clonotypes.

## Discussion

Herein, we characterised venom-specific T cell responses during the first 6 months of VIT. Although it cannot be used solely to determine clinical reactivity, specific IgG4 during VIT is considered as a marker of treatment success and tolerance development [5,23,24]. Accordingly, our data indicate an early (7 days) venom- and Ves v 5-specific IgG4 increase after VIT initiation, actually earlier than previously reported for ultra-rush VIT [7,25], suggesting an activation of pre-existing allergen-specific B cells. This was accompanied by a transient increase in specific IgE at d7, which returned to baseline by mo6, resulting in an increase in the specific IgG4/IgE ratio only at mo6.

Older reports on venom-specific T-cell responses during VIT had a methodologically limited resolution of individual T-cell subsets [6,26] and described phenotypic changes in the broader Th2 compartment lacked allergen specificity, which impeded the identification of disease-specific signatures [7]. Therefore, we characterised venom-specific T cells by single-cell RNA sequencing for deep phenotyping. We identified and isolated venom-specific T cells after stimulation with native venom extract followed by the detection of activation markers. Compared to tetramer-based approaches, this strategy captures a broader allergen-specific T-cell compartment without HLA restriction but may underestimate anergic cells that fail to respond to stimulation. At baseline, venom-specific T cells comprised Tregs, multiple Th2 populations, and subsets expressing XCL1, IL2, or CCL20 and IL4I1. Expression of IL4, IL13, and IL5 was consistent with previous reports [6], while, to our knowledge, IL31 and granzyme B expression in venom-specific Th2 cells has not previously been described. Granzyme B expression has previously been reported in Th2 [27] and shown to be upregulated by IL-10 [28], but its weak correlation with baseline Ves v 5-specific IgE and no apparent association with tolerance induction suggest a limited role in the treatment response.

Notably, IL5⁺IL3^-^ T cells, corresponding to a terminally differentiated Th2 subset [22], strongly correlated with baseline Ves v 5-specific IgE levels, supporting their possible functional role and their potential as a biomarker. In addition, the frequency of IL5⁺IL3^-^ T cells increased during VIT. As this increase correlated with the Ves v 5-specific IgG4/IgE ratio at mo6, one can speculate that IL-5 contributes to the induction of IgG4. Although IL-5 has not been directly implicated in IgG4 production, it promotes B-cell proliferation and differentiation [29,30] and could act alongside IL-4 and IL-10, both established drivers of IgG4-producing B-cell differentiation [31].

IL5⁺IL3^-^ Th2 cells also exhibited the strongest expression of exhaustion markers during VIT, which was associated with a stronger IgG4 response. We suppose that this phenotype may be a consequence of the repetitive antigen stimulation during VIT. Notably, we did not observe substantial changes in expression of the activation markers HLA-DR or CD38, or the proliferation marker MKI67, although this could be due to a suboptimal sampling time. This suggests that the increased expression of exhaustion markers reflects a genuine exhaustion phenotype rather than a transient consequence of recent *in vivo* activation. Indeed, although exhaustion markers may be transiently upregulated following activation, their sustained expression in the absence of concomitant increases in activation or proliferation markers is more consistent with a state of T-cell exhaustion. However, this finding will need to be experimentally validated. The association between increased CTLA4 [32] and PD-1 [33] expression and IgG4-related disease further support a potential link between T-cell exhaustion and IgG4-associated immune regulation, although these markers were not reported on the same T-cell populations.

TCR repertoire analysis revealed the absence of clonotypes shared between individuals but substantial persistence across time points. Clonal expansion and deletion were limited, whereas transitions between T-cell clusters were frequent, particularly among Th2 populations. Together with the observed phenotypic changes, these findings suggest that VIT is associated with phenotypic reprogramming of pre-existing venom-specific T-cell clonotypes rather than extensive clonal expansion.

We identified two Treg clusters distinguished by differential expression of LGALS3 and HLA-DP/DR, which we attributed to differences in activation and potential suppressive function. LGALS3 expression was of particular interest, given its association with IgG4 responses in IL10-producing Tfh cells [34]. We did not detect IL10 or TGFB1 transcripts, but this should not be interpreted as evidence of their absence, as low-abundance transcripts may be missed by scRNAseq [35]. The lack of distinct markers separating both subsets hindered their isolation for functional characterisation. The overall frequency of venom-specific Tregs negatively correlated with Ves v 5-specific IgE at d0. Consistent with previous findings [36], the proportion of venom-specific Tregs decreased during VIT, potentially reflecting migration to tissues. However, the remaining Tregs displayed increased activation, which strongly correlated with the increase in the Ves v 5-specific IgG4/IgE ratio at mo6. TCR analysis revealed a highly polyclonal repertoire, suggesting a response to a broad range of antigens, with the emergence of new clones during VIT. Moreover, the lack of TCR overlap between Tregs and memory Tconv argues against differentiation of Tconv into Tregs. Together, these findings suggest that VIT promotes the activation and potential functional maturation of venom-specific Tregs, which may contribute to IgG4-associated tolerance.

We did not detect venom-specific CD8 T cells above background. Although a type 2 CD8 T-cell (TC2) population expressing GATA3 and type 2 cytokines has been implicated in allergy [37], these cells are predominantly found in tissues and may therefore be present below the detection limit in peripheral blood.

This study has several limitations. The use of scRNAseq enabled high-resolution characterisation of immune cell phenotypes and antigen receptor repertoires, but at the cost of limited throughput – although enriching populations of interest and multiplexing samples enabled analysis of 32 samples with sufficient resolution for comparative analyses. However, the absence of an allergic control group not receiving VIT prevented assessment of natural temporal variations, particularly in antigen receptor repertoires. In addition, we focused exclusively on wasp venom allergy, and the generalisability of these findings to other allergic diseases remains to be established.

In conclusion, we characterised the venom-specific T cell repertoire in wasp venom allergy at high resolution and identified IL5⁺IL3⁻ CD4 T cells as a population associated with elevated Ves V 5-specific IgE. VIT induced marked changes in the venom-specific CD4 T-cell compartment, particularly regarding IL5⁺IL3⁻ CD4 T cells and Tregs, and these changes correlated with desensitisation as assessed by the specific IgG4/IgE ratio. Notably, several changes were detectable as early as 7 days after VIT initiation, identifying potential candidates for early biomarkers of treatment response. The association of IL5⁺IL3⁻ CD4 T cells with both allergic sensitisation and VIT-induced immune modulation suggest that IL5-related pathways may contribute to the response to allergen immunotherapy.

---

**Table S1:** Summary of cohorts.

| **Cohorts** | **Non-allergic<br>(n=5)** | **Venom-allergic<br>d0 (n=9)** | **Venom-allergic<br>d7 (n=9)** | **Venom-allergic<br>mo6 (n=9)** |
|---|---|---|---|---|
| Male (prop) | 1.00 | 0.33 | 0.33 | 0.33 |
| Age in year (mean, SD) | 27.8 (4.38) | 53.3 (9.23) | 53.3 (9.23) | 53.3 (9.23) |
| BMI (mean, SD) | 24.8 (2.45) | 27.1 (6.34) | 27.1 (6.34) | 27.1 (6.34) |
| Total IgE (mean, SD) | 26.7 (30.5) | 233.5 (188.9) | 261.6 (202.0) | 209.9 (194.4) |
| Venom IgE (mean, SD) | 0.22 (0.13) | 19.52 (27.94) | 25.47 (31.25) | 14.44 (18.35) |
| Ves v 1 IgE (mean, SD) | 0.10 (0.01) | 4.08 (10.54) | 8.44 (13.96) | 3.61 (4.35) |
| Ves v 5 IgE (mean, SD) | 0.16 (0.11) | 16.28 (32.51) | 24.32 (31.77) | 11.27 (16.22) |
| Venom IgG4 (mean, SD) | 0.07 (0.03) | 0.86 (0.79) | 4.17 (2.56) | 5.70 (5.56) |
| Ves v 1 IgG4 (mean, SD) | 0.11 (0.04) | 0.34 (0.43) | 1.16 (1.76) | 2.47 (3.79) |
| Ves v 5 IgG4 (mean, SD) | 0.04 (0.03) | 0.55 (0.70) | 3.00 (1.76) | 2.90 (2.00) |

**Table S2:** BAT antibody panel.

| **Staining** | **Marker** | **Fluorophore** | **Clone** | **Manufacturer** |
|---|---|---|---|---|
| Surface | CD203c | BV421 | NP4D6 | Biolegend |
| Surface | CD123 | BV605 | 6H6 | Biolegend |
| Surface | HLADR | Percp | L243 | Biolegend |
| Surface | CD63 | APC-Cy7 | H5C6 | Biolegend |
| Surface | CD33 | BV510 | WM53 | Biolegend |
| Surface | CD300a | PE | MEM-260 | Invitrogen |
| Surface | CD32b | PE-Cy7 | S18005H | Biolegend |
| Surface | CD62L | Alexa Fluor 700 | DREG-56 | Biolegend |
| Surface | Live/Dead | Zombie yellow |  | Biolegend |
| Intracellular | SYK | Alexa Fluor 488 | 4D10.1 | eBioscience |
| Intracellular | S6 p265/236 | APC | REA454 | Miltenyi biotech |

**Table S3:** venom-specific T cell antibody panel.

| **Conjugate** | **Marker** | **Fluorophore** | **Clone** | **Manufacturer** |
|---|---|---|---|---|
|  | Live/Dead | Blue |  | Invitrogen |
| Fluorochrome | CD25 | VB423 | REA945 \| 4E3 | Miltenyi |
| Fluorochrome | CD45RA | Viogreen | REA562 \| T6D11 | Miltenyi |
| Fluorochrome | CD127 | VB600 | REA614 \| A019D5 | Miltenyi |
| Fluorochrome | CD4 | VB646 | REA623 \| SK3 | Miltenyi |
| Fluorochrome | CD137 | PE | REA765 \| 4B4-1 | Miltenyi |
| Fluorochrome | CD27 | Percp-Vio700 | REA499 \| LG.3A10 | Miltenyi |
| Fluorochrome | CD154 | APC | REA238 \| 5C8 | Miltenyi |
| Fluorochrome | CD8 | R700 | REA734 \| HIT8a | Miltenyi |
| Oligonucleotide | CD3 | Totalseq-C | UCHT1 | Biolegend |
| Oligonucleotide | NOTCH2 | Totalseq-C | MHN2-25 | Biolegend |
| Oligonucleotide | Hashtag | Totalseq-C | LNH-94; 2M2 | Biolegend |

**Table S4:** venom-specific T cell clusters.

| Cluster | Population | Genes | CD3E protein | Notch 2 protein |
|---|---|---|---|---|
| 8 | CD8 Granzyme<sup>+</sup> | CD8A+ TNFRSF9<sup>+</sup> GZMB<sup>+</sup> | / | / |
| 12 | Low-quality cells | CD4+ NFAT<sup>high</sup> | / | / |
| 1 | Treg act low | CD4<sup>+</sup> TNFRSF9<sup>+</sup> CD40LG<sup>-</sup> FOXP3<sup>+</sup> LGALS3<sup>low</sup> HLA-DPA1/DPB1<sup>low</sup> HLA-DRA<sup>low</sup> | Mid | Mid |
| 3 | Treg act high | CD4<sup>+</sup> TNFRSF9<sup>+</sup> CD40LG<sup>-</sup> FOXP3<sup>+</sup> LGALS3<sup>high</sup> HLA-DPA1/DPB1<sup>high</sup> HLA-DRA<sup>high</sup> | Up | Mid |
| 2 | Th2 no cytokines | CD4<sup>+</sup> TNFRSF9<sup>+</sup> CD40LG<sup>+</sup> GATA3<sup>low</sup> IL4<sup>-</sup> IL13<sup>-</sup> IL3<sup>-</sup> | Mid | Mid |
| 5 | Th2 classical | CD4<sup>+</sup> TNFRSF9<sup>+</sup> CD40LG<sup>+</sup> GATA3<sup>+</sup> IL4<sup>+</sup> IL13<sup>+</sup> IL3<sup>+</sup> | Down | Mid |
| 6 | Th2 Granzyme<sup>+</sup> | CD4<sup>+</sup> TNFRSF9<sup>+</sup> CD40LG<sup>+</sup> GATA3<sup>+</sup> IL4<sup>+</sup> IL13<sup>+</sup> IL3<sup>+</sup> GZMB<sup>+</sup> | Down | Up |
| 7 | Th2 IL31<sup>+</sup> | CD4<sup>+</sup> TNFRSF9<sup>+</sup> CD40LG<sup>+</sup> GATA3<sup>+</sup> IL4<sup>+</sup> IL13<sup>+</sup> IL3<sup>+</sup> IL31<sup>+</sup> | Down | Mid |
| 9 | Th2 IL5<sup>+</sup> IL3<sup>-</sup> | CD4<sup>+</sup> TNFRSF9<sup>+</sup> CD40LG<sup>+</sup> GATA3<sup>+</sup> IL4<sup>+</sup> IL13<sup>+</sup> IL3<sup>-</sup> IL5<sup>+</sup> | Mid | Mid |
| 4 | CD4 CCL20<sup>+</sup> IL4l1<sup>+</sup> | CD4+ TNFRSF9<sup>+</sup> CD40LG<sup>+</sup> CCL20<sup>+</sup> IL4l1<sup>+</sup> LGALS3<sup>+</sup> | Mid | Up |
| 10 | CD4 XCL1<sup>+</sup> | CD4+ TNFRSF9<sup>+</sup> CD40LG<sup>+</sup> XCL1<sup>+</sup> | Down | Mid |
| 11 | CD4 IL2<sup>+</sup> | CD4+ TNFRSF9<sup>+</sup> CD40LG<sup>+</sup> IL2<sup>high</sup> | Down | Mid |

**Supplementary Figure S1:**
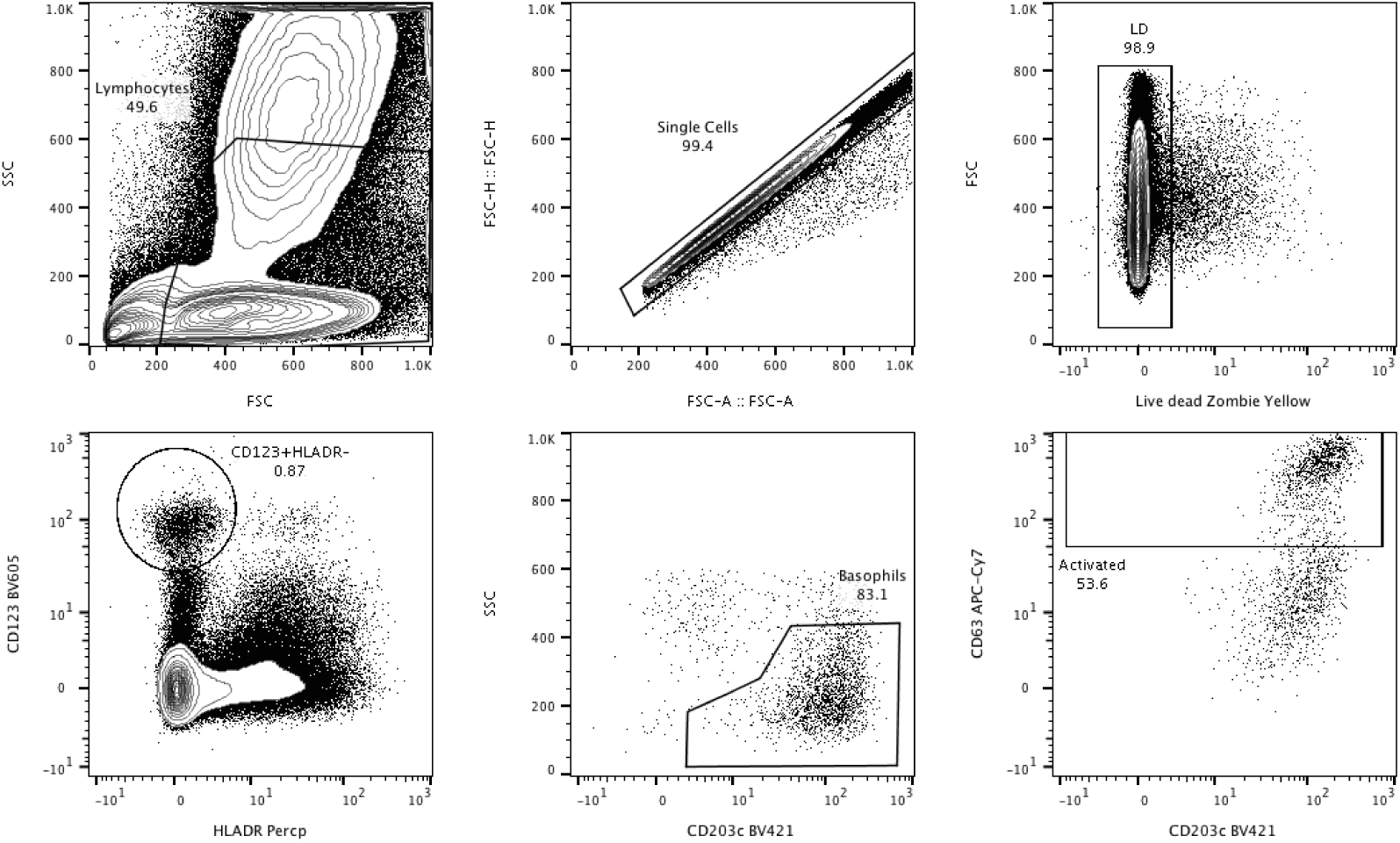
Gating strategy of the Basophil activation test.

**Supplementary Figure S2:**
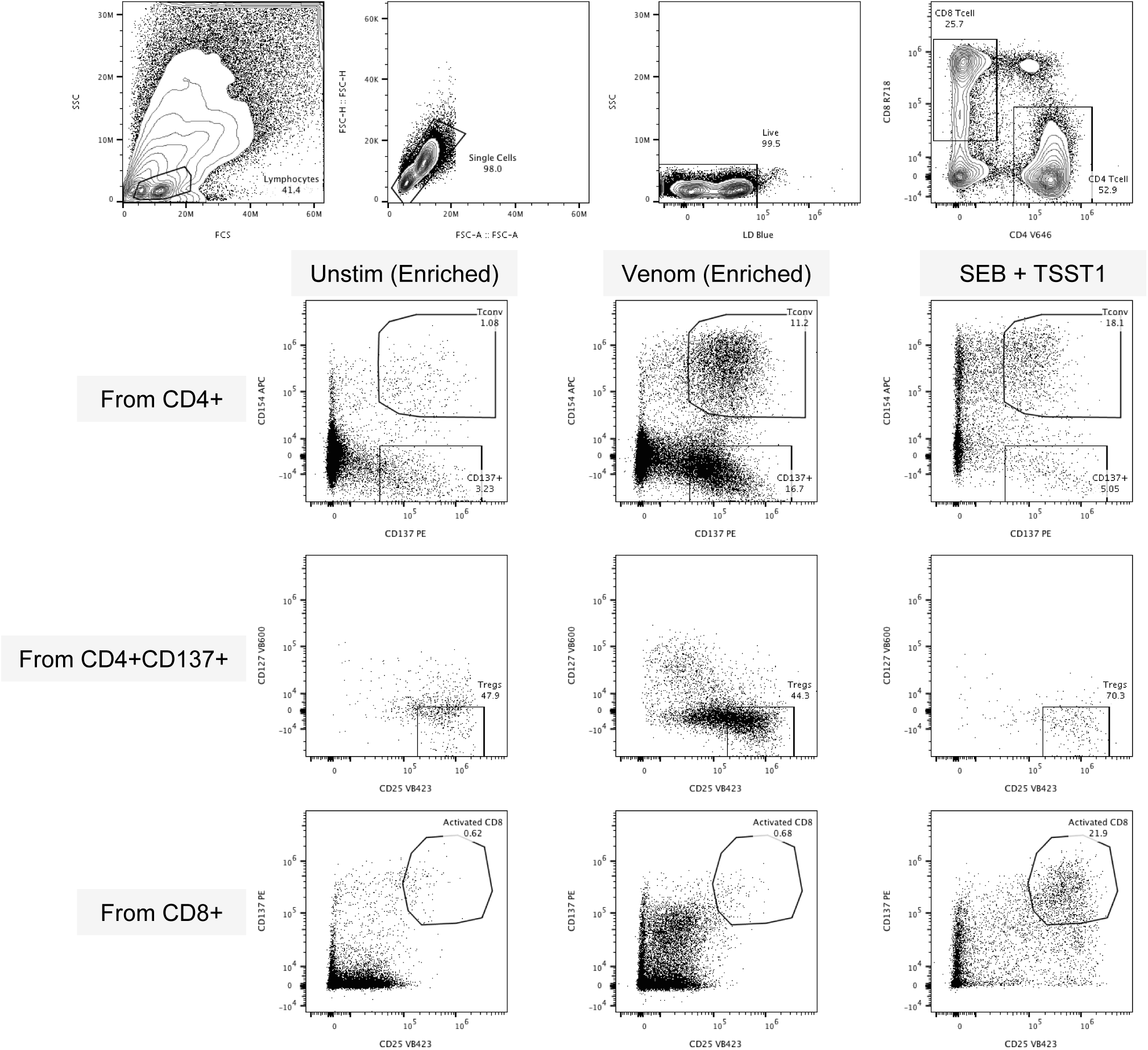
Gating strategy for sorting of the venom-specific T cells.

**Supplementary Figure S3:**
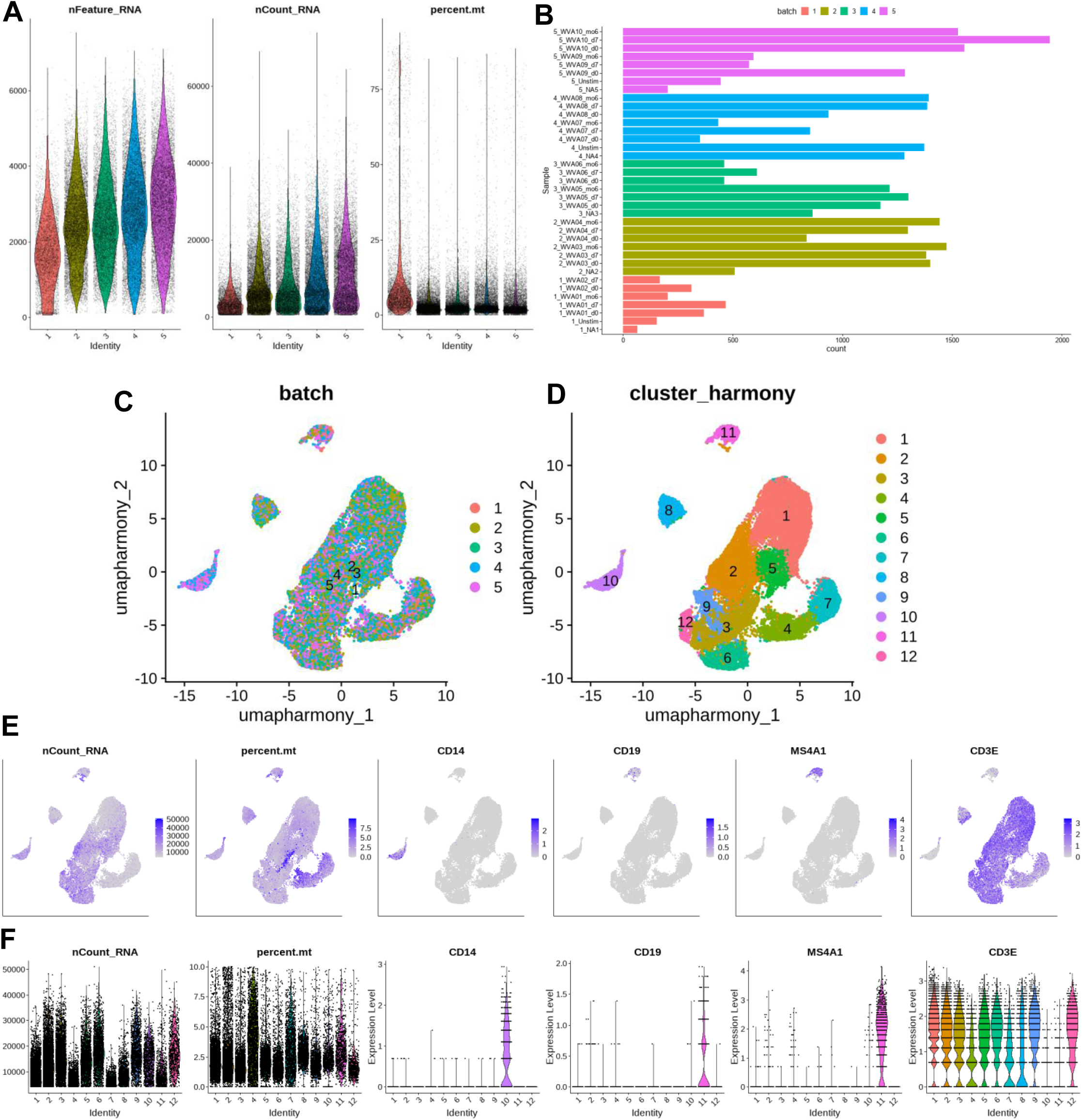
Quality control, integration and first clustering of the scRNAseq dataset using Seurat v5. **A:** Number of features, RNA and percentage of mitochondrial genes in each of the sequenced cells. **B:** Number of cells per sample after quality control filtering. **C:** UMAP depicting batches after integration and batch correction and **D:** clusters obtained using Leiden clustering. **E,F:** Statistical parameters and selected features overlaid on **E:** UMAP and **F:** as boxplot per cluster.

**Supplementary Figure S4:**
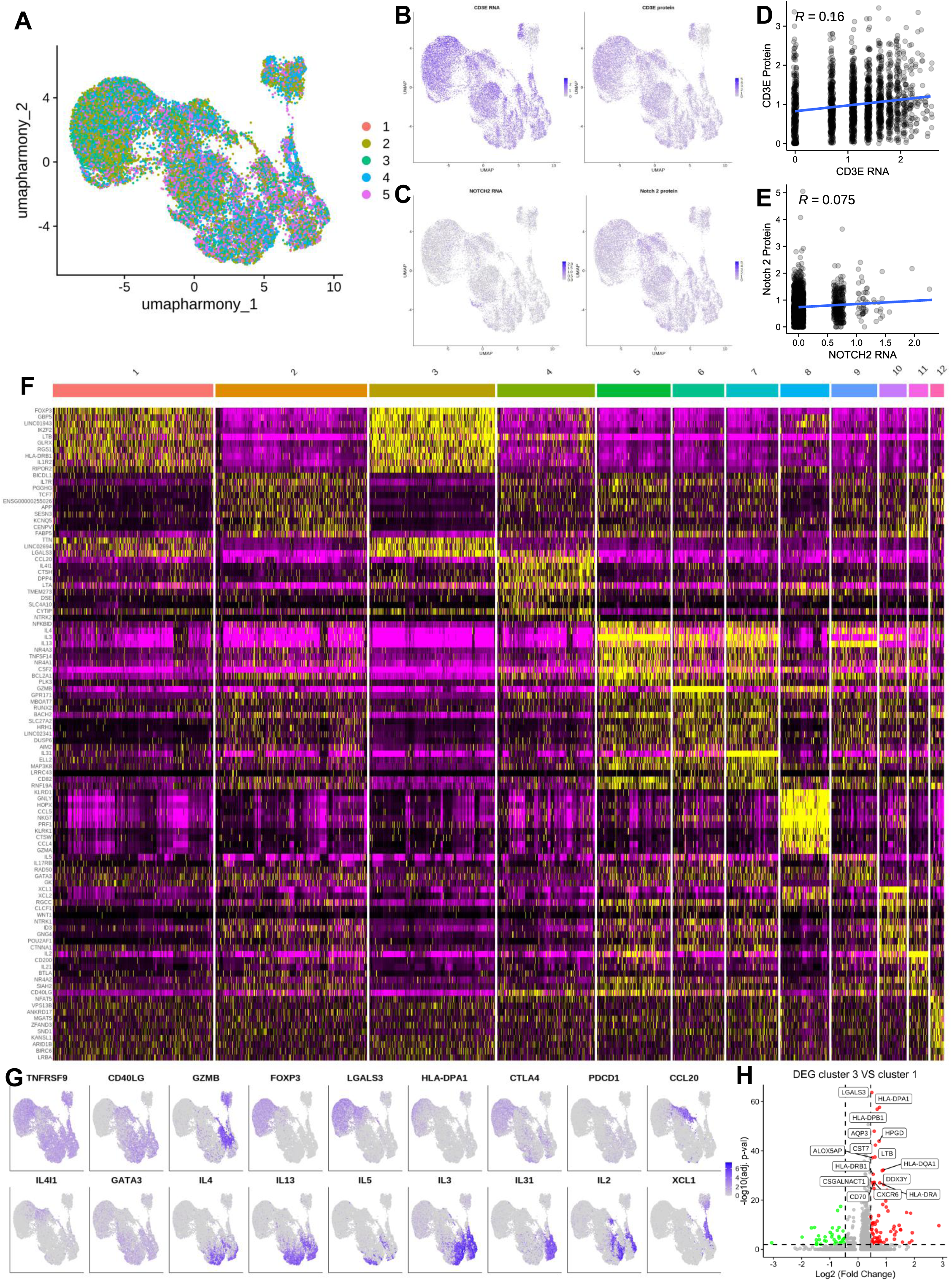
T cells integration and clustering. **A-C:** Umap depicting **A:** batches after correction, RNA and protein expression of **B:** CD3e and **C:** Notch2. **D-E:** Correlation between RNA and protein expression of **D:** CD3e and **E:** Notch2. **F:** Heatmap displaying top differentially expressed genes in T cell clusters. **G:** Selected RNA expression displayed on UMAPs. **H:** Volcano plot depicting differentially expressed genes between clusters 3 and 1.

**Supplementary Figure S5:**
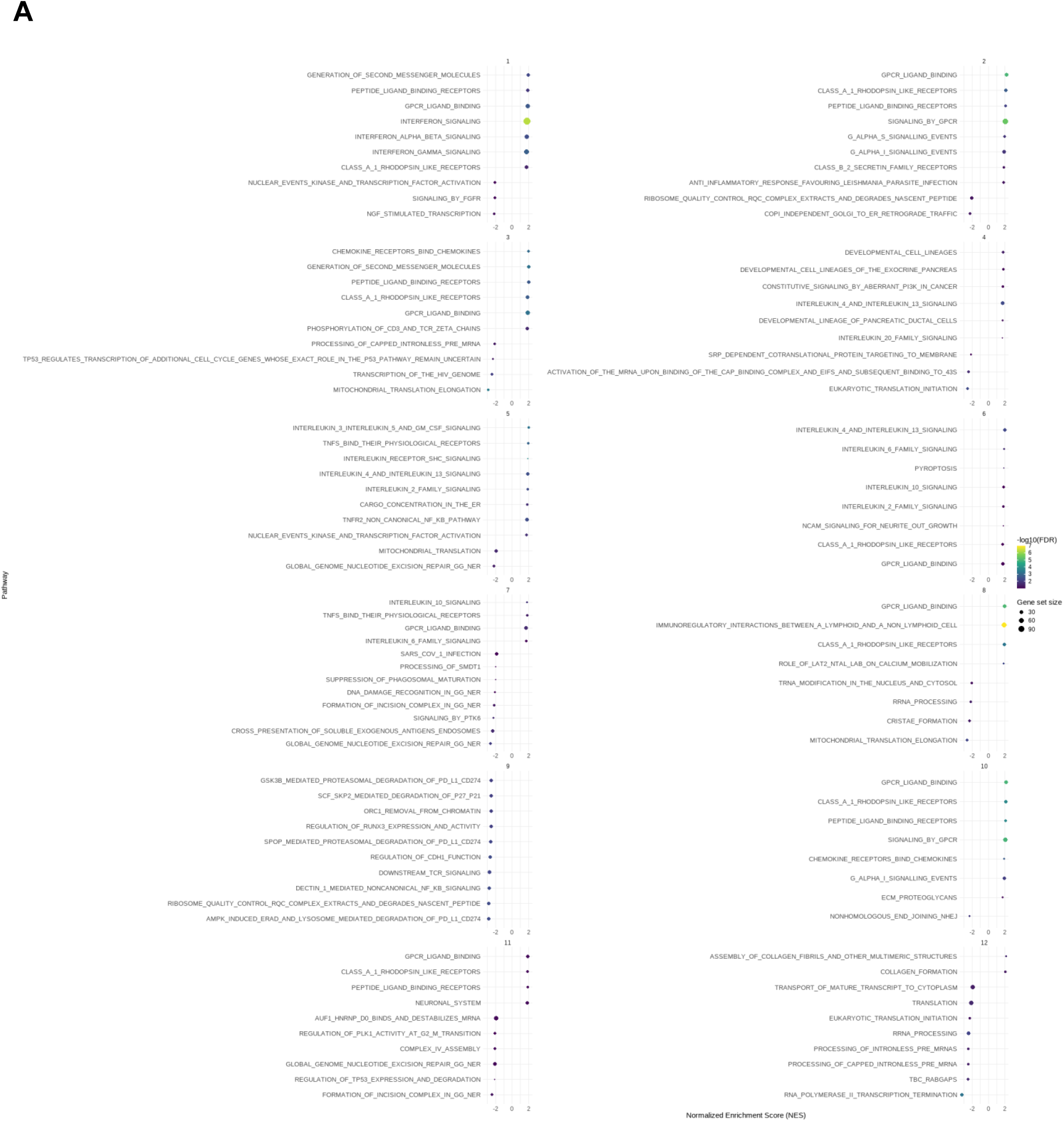
Gene set enrichment analysis of venom-specific T cells using the Reactome database.

**Supplementary Figure S6:**
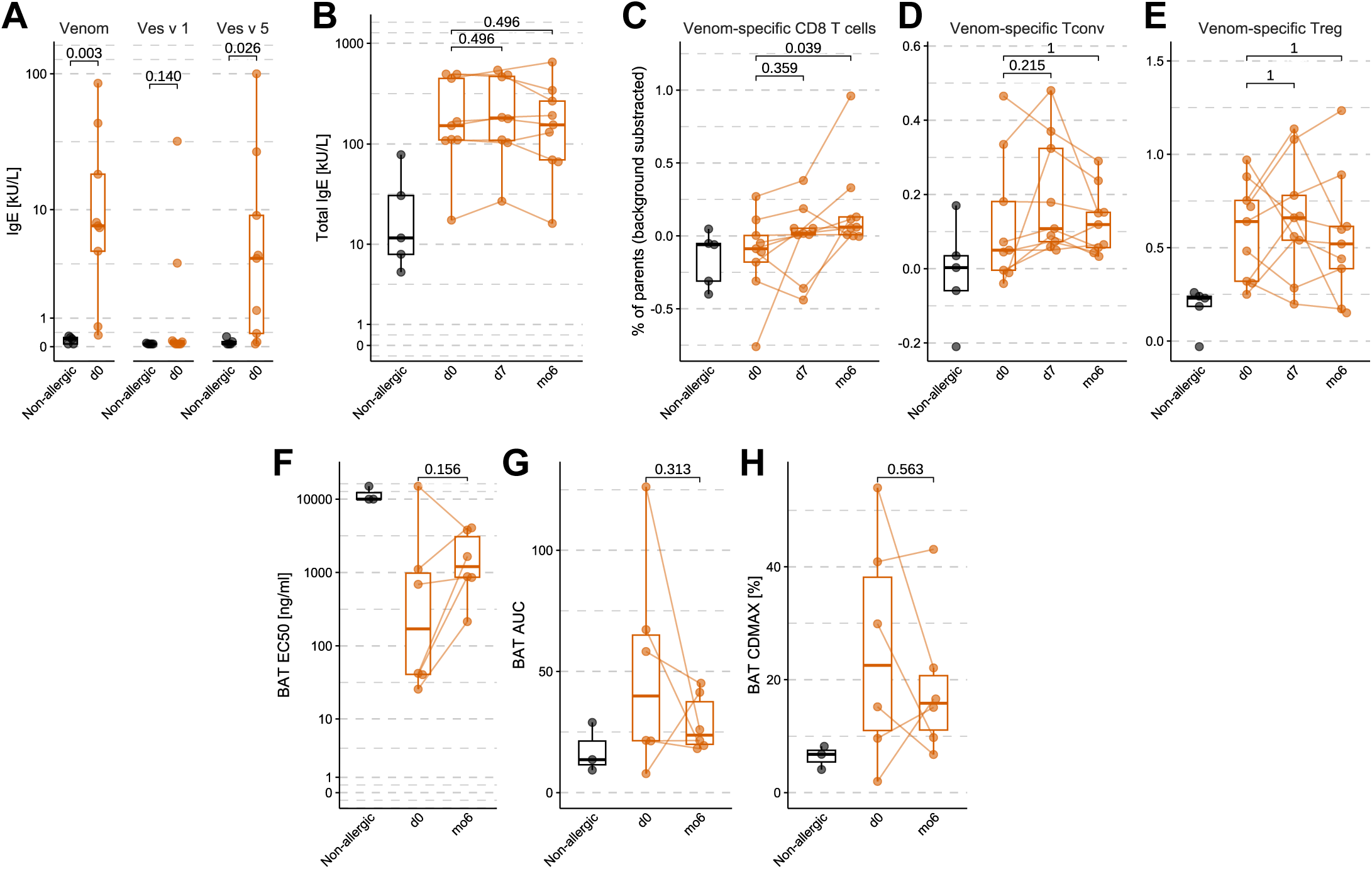
**A:** Venom, Ves v 1 and Ves v 5 IgE levels [kU/L] and **B:** Total IgE level [kU/L] in non-allergic (black, n=5) and venom-allergic (orange, n=9). **C:** Percentage of venom-specific CD8, **D:** venom-specific CD4 Tconv and **E:** venom-specific Treg measured by flow cytometry. **F:** EC50 [ng/ml], **G:** Area under the curve (AUC) and **H:** Maximum of activated basophil of the BAT. Statistical tests were conducted using the Wilcoxon signed-rank paired test corrected for multiple comparisons between time points with Benjamini-Hochberg (FDR).

**Supplementary Figure S7:**
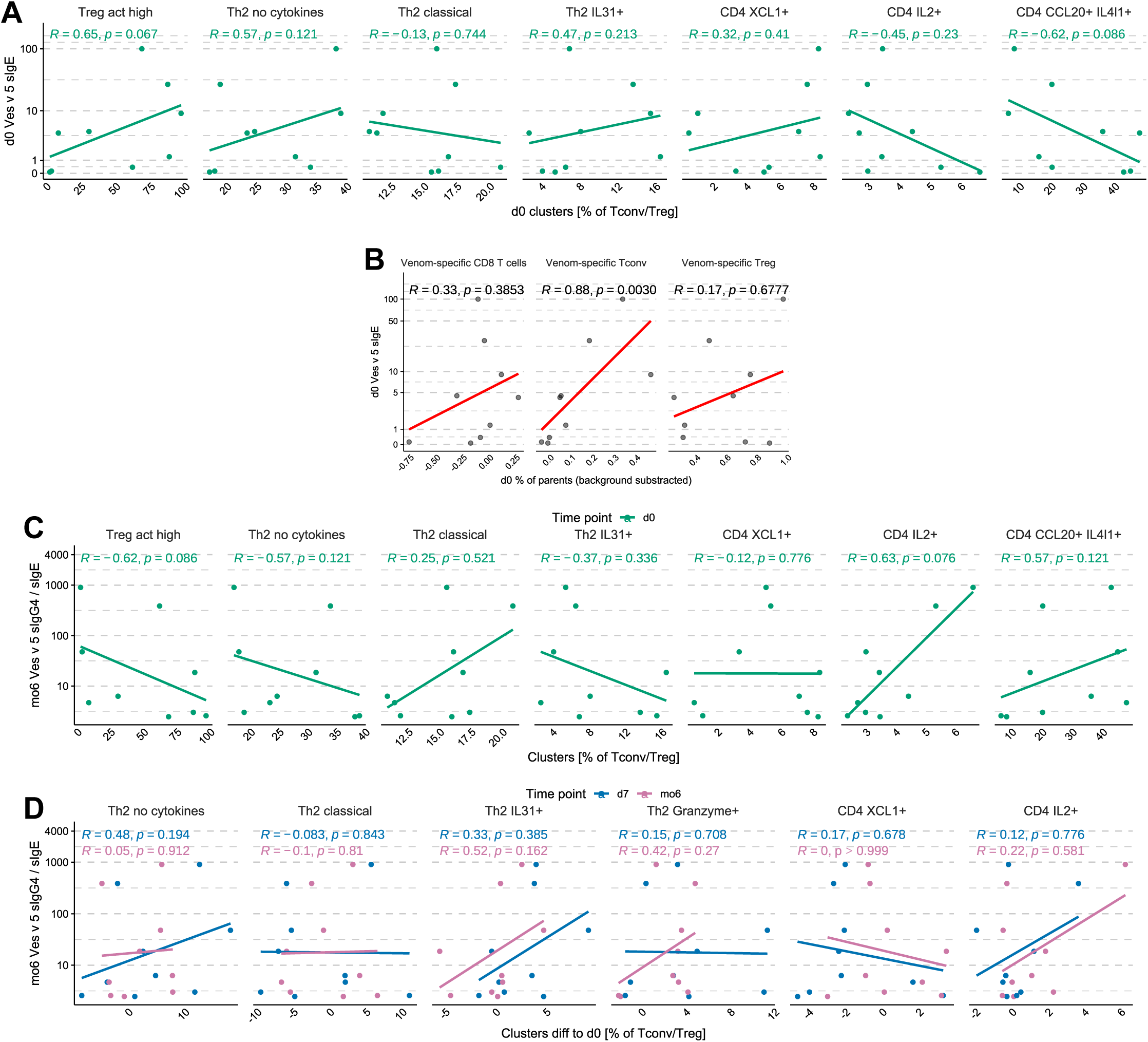
**A:** Spearman correlation of venom-specific T cell populations percentages determined using scRNAseq with Ves v 5-specific IgE in venom-allergic individuals at d0. **B:** Spearman correlation of venom-specific CD8 T cell, Tconv or Treg percentages with Ves v 5-specific IgE in venom-allergic individuals at d0. **C:** Spearman correlation of venom-specific T cell populations percentages determined using scRNAseq at d0 with Ves v 5-specific IgG4 / IgE ratio in venom-allergic individuals at mo6. **D:** Spearman correlation of venom-specific T cell populations percentages at d7 and mo6, normalised to d0 with Ves v 5-specific IgG4 / IgE ratio in venom-allergic individuals at mo6.

**Supplementary Figure S8:**
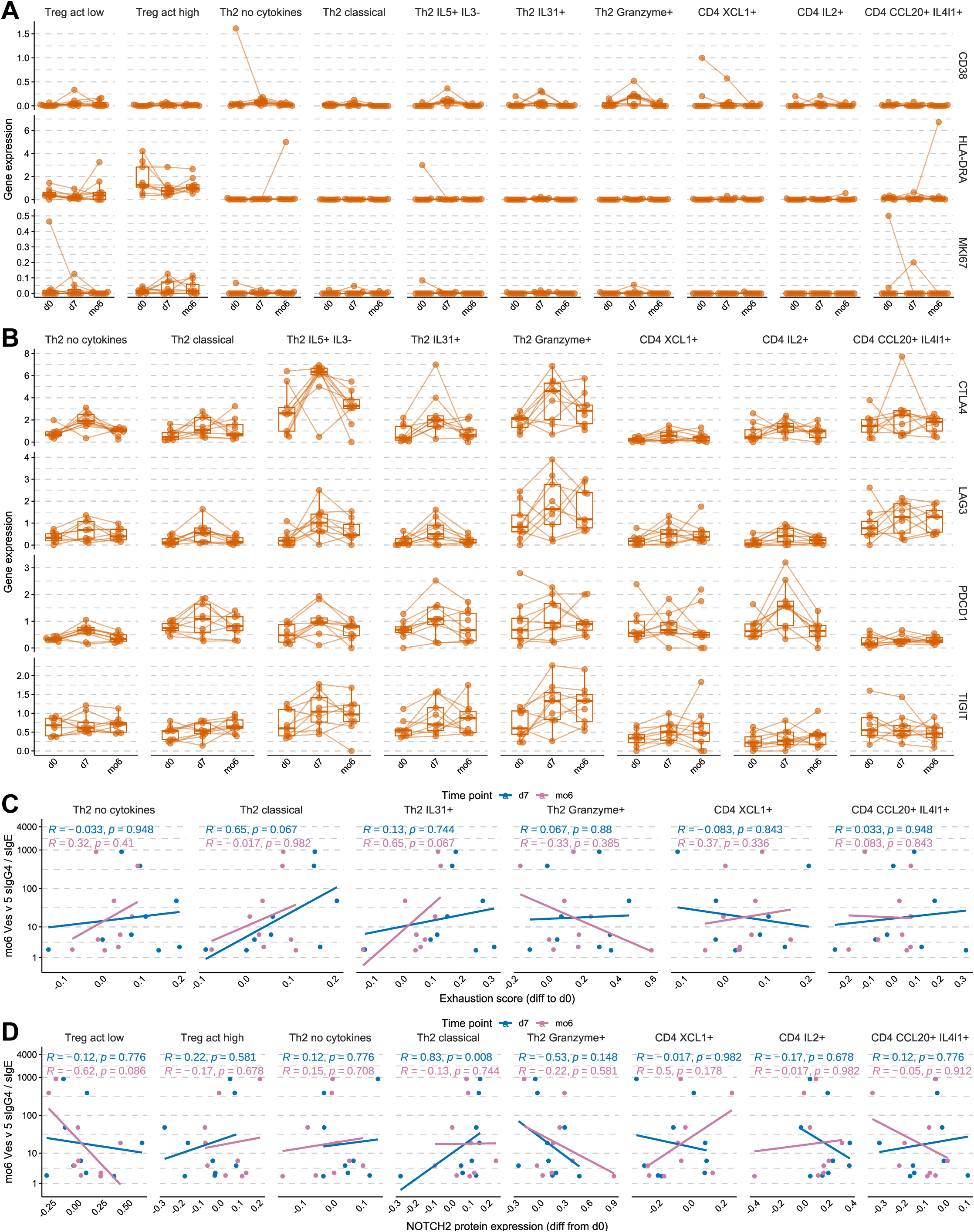
**A-B:** Boxplots depicting gene expression of selected genes in venom-allergic at d0, d7 and mo6 (n=9). C: Spearman correlation of the exhaustion score normalised to d0 in venom-specific T cell populations at d7 and mo6 with Ves v 5-specific IgG4 / IgE ratio in venom-allergic individuals at mo6. D: Spearman correlation of the Notch2 protein expression normalised to d0 in venom-specific T cell populations at d7 and mo6 with Ves v 5-specific IgG4 / IgE ratio in venom-allergic individuals at mo6.

**Supplementary Figure S9:**
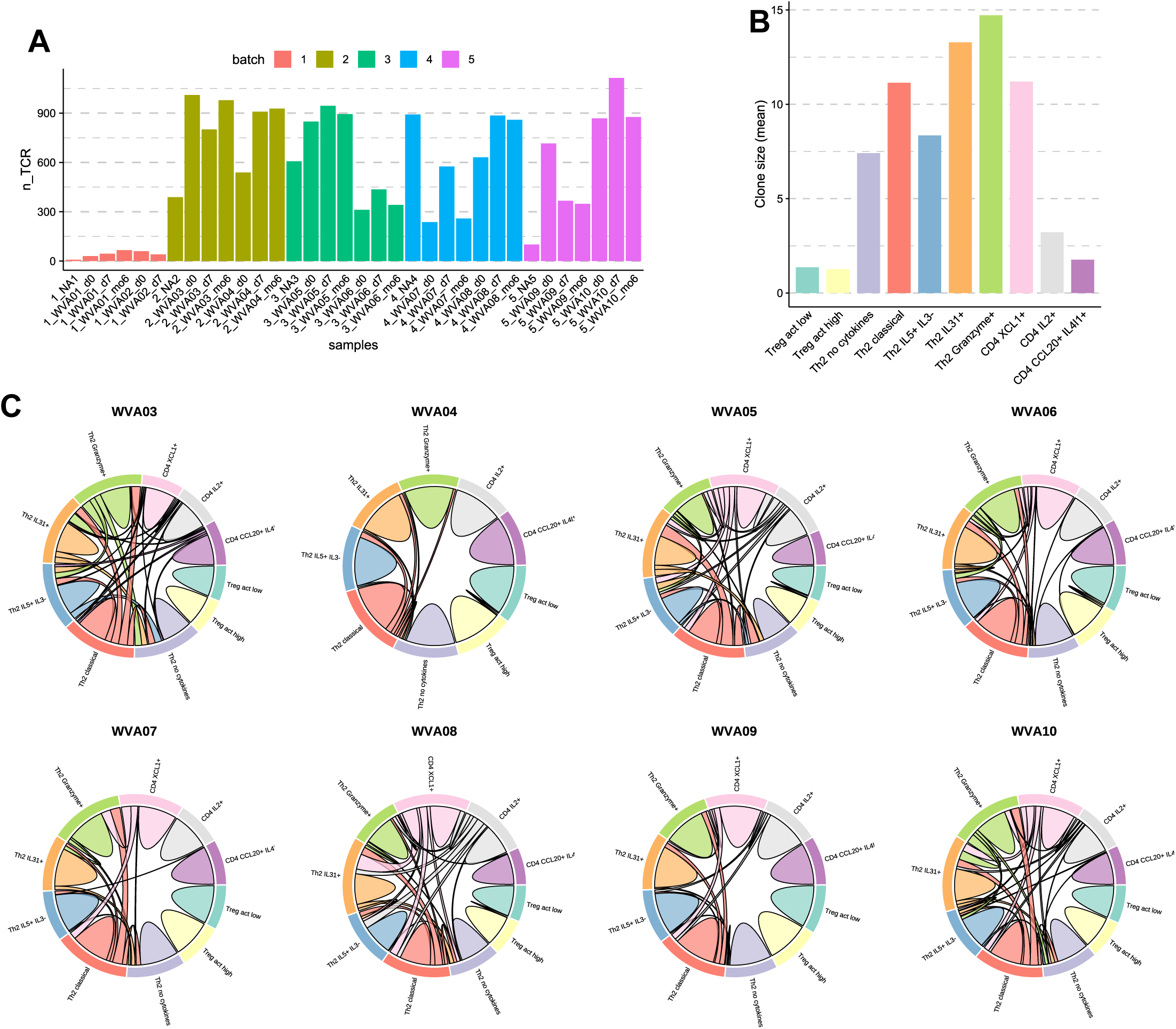
TCR repertoire clonality analysis. **A:** Number of sequenced TCR per sample. **B:** Mean clone size per venom-specific T cell cluster. **C:** Circos plot showing clonotypes shared by multiple T cell populations at d0 separated by venom-allergic.

**Supplementary Figure 10:**
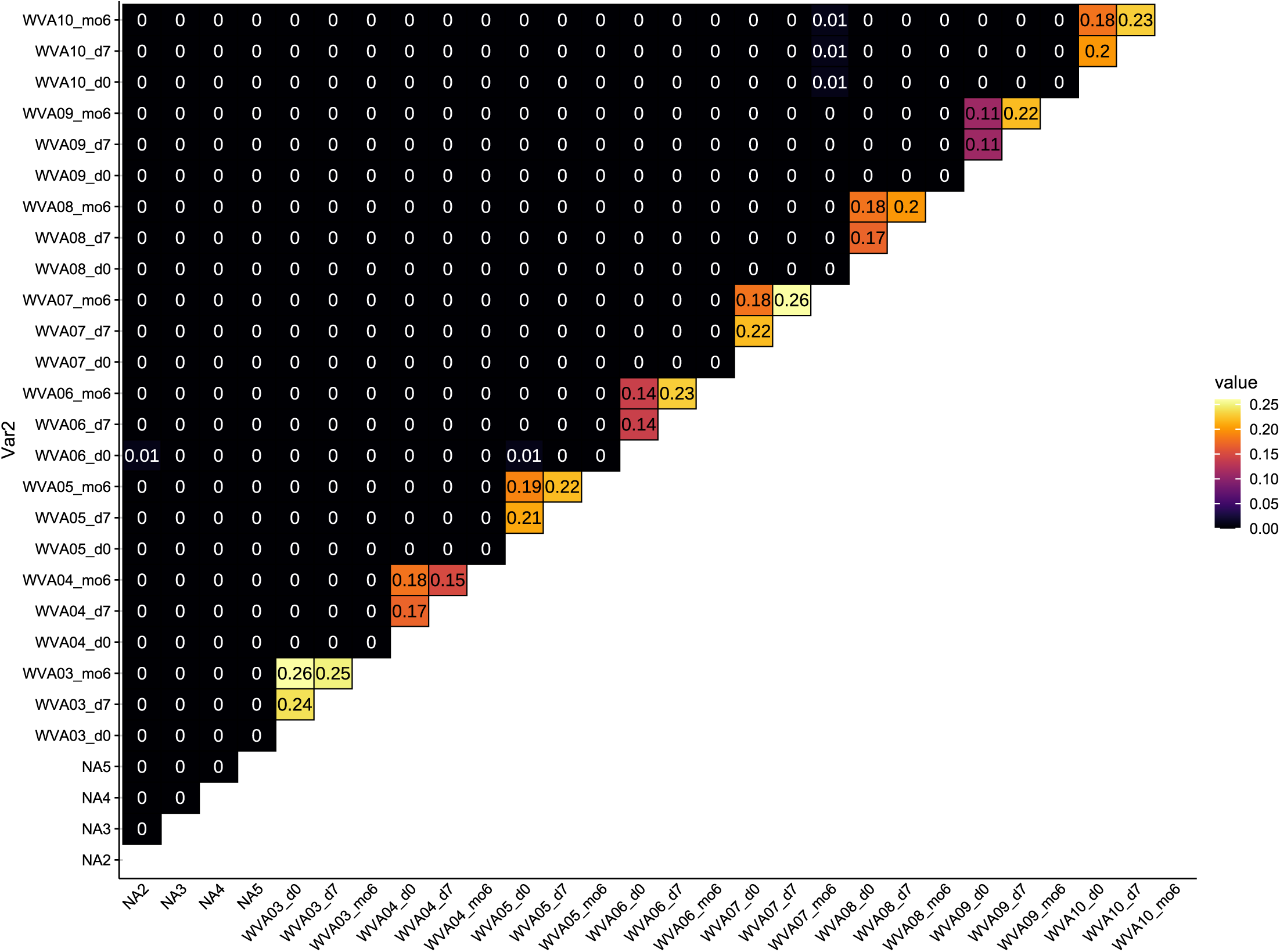
TCR repertoire analysis. TCR repertoire overlap between donor and time points.

**Supplementary Figure 11:**
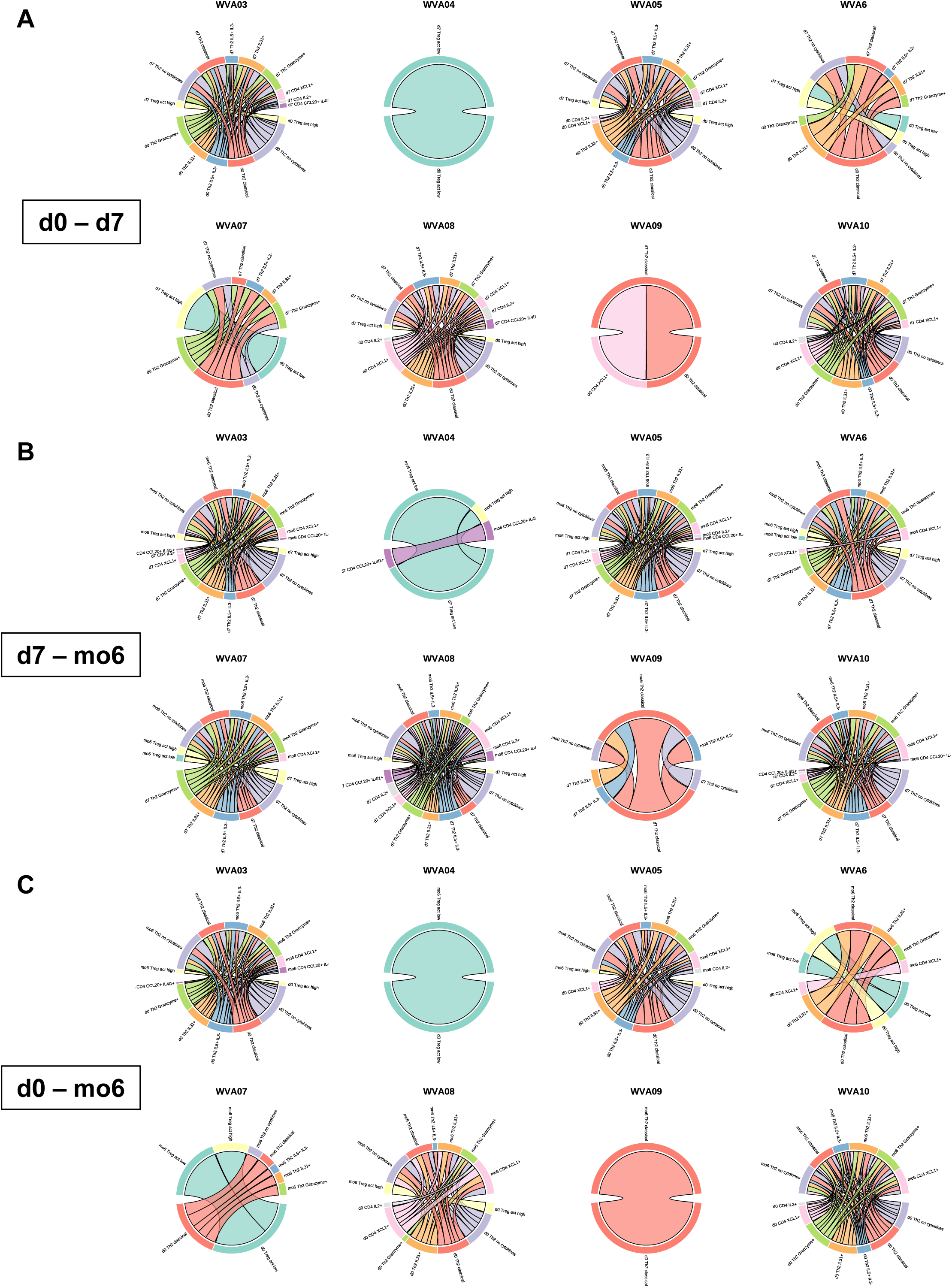
TCR repertoire analysis. **A-C:** Circos plots depicting TCR clones transitioning between venom-specific T cell populations and indicated time points.

**Supplementary Figure 12:**
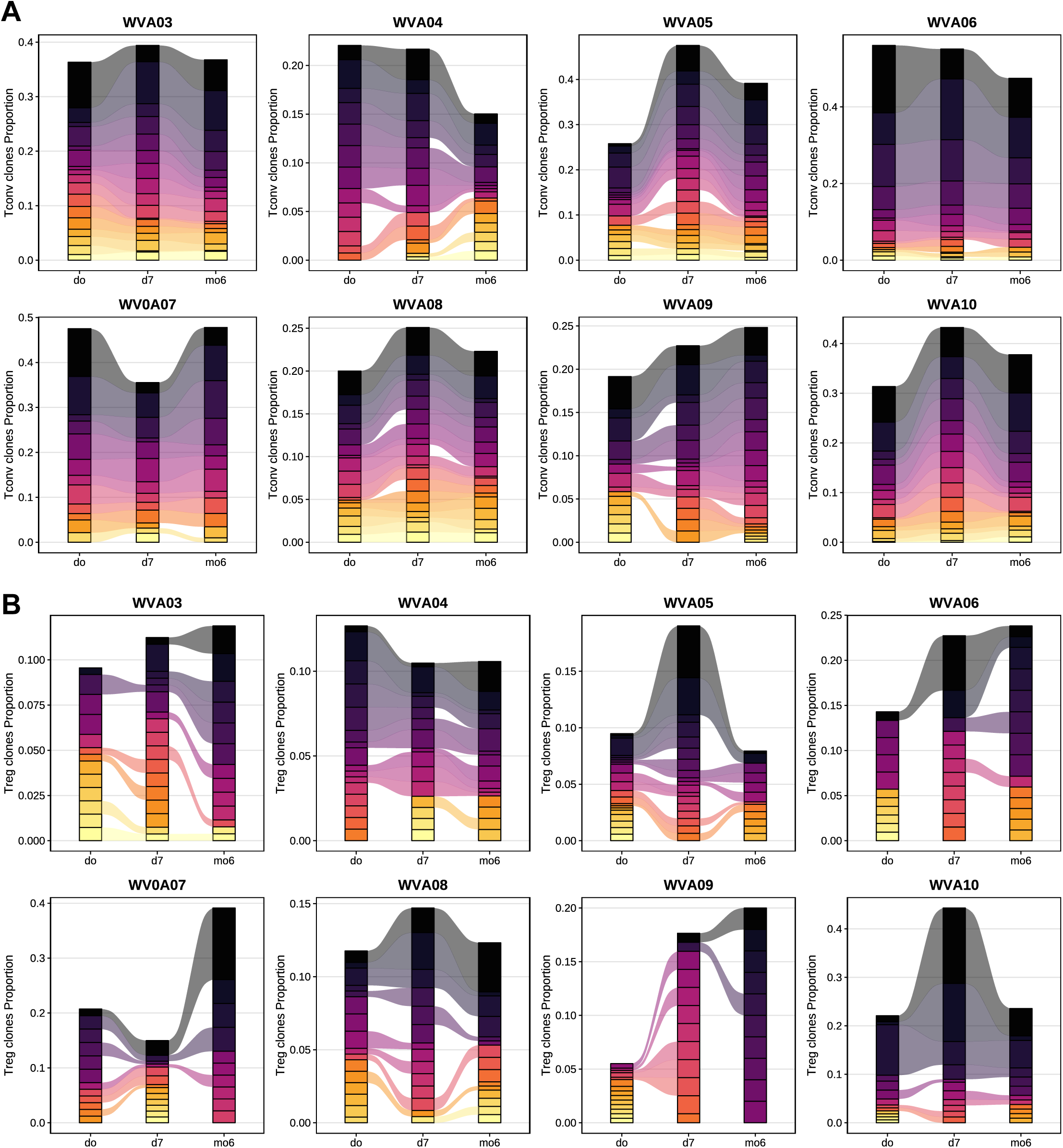
TCR repertoire analysis. **A-B:** Plot depicting during VIT the dynamics of top clones for each venom-allergic in **A:** venom-specific Tconv and **B:** venom-specific Tregs.

## Notes

### Competing Interest Statement

The authors have declared no competing interest.

### Author Declarations

Ethics committee (EA2/304/21) of Charite - Universitatsmedizin Berlin gave ethical approval for this work.

